# Polygenic and machine learning analysis of medication response in a prospective pediatric cohort initiating ADHD medication

**DOI:** 10.64898/2026.09.09.26362635

**Authors:** Alba Escalera-Balsera, Maria M Lilja, Mattias Nordstrand, Terje Falck-Ytter, Eva Serlachius, Jyoti Bhagia, Kristiina Tammimies, Linda Halldner

**Affiliations:** Center of Neurodevelopmental Disorders (KIND), Centre for Psychiatry Research, Department of Women’s and Children’s Health, Karolinska Institutet, Region Stockholm, Stockholm, Sweden; Astrid Lindgren Children’s Hospital, Karolinska University Hospital, Region Stockholm, Stockholm, Sweden; Department of Clinical Sciences, Child and Adolescent Psychiatry, Umea University, Umea, Sweden; Development and Neurodiversity Lab (DIVE), Department of Psychology, Uppsala University, Uppsala, Sweden; Center of Neurodevelopmental Disorders (KIND), Centre for Psychiatry Research, Department of Women’s and Children’s Health, Karolinska Institutet and Stockholm Health Care Services, Region Stockholm, Stockholm, Sweden; Centre for Psychiatry Research, Department of Clinical Neuroscience, Karolinska Institutet, Region Stockholm, Stockholm, Sweden; Department of Clinical Sciences, Lund, Lund University, Lund, Sweden; Department of Psychiatry and Psychology, Mayo Clinic, Rochester, MN, USA; Department of Medical Epidemiology and Biostatistics, Karolinska Institutet, Stockholm, Sweden

**Keywords:** Pediatric ADHD, medication response, polygenic scores, educational attainment, machine learning

## Abstract

While medication for attention deficit hyperactivity disorder (ADHD) is effective, predictors of treatment response remain poorly understood. Given the use of polygenic scores (PGS) in psychiatric conditions, we investigate their potential to predict ADHD medication response. In this work, we recruited 309 participants aged 6-17 years from the Swedish ADHD medication and predictors of treatment outcome (ADAPT) study (ClinicalTrials.gov: NCT02136147; June 2015), and calculated their PGS. Compared with a reference cohort (N=780), ADAPT showed higher PGS for ADHD (*p*<2.22e-16), and lower PGS for autism (*p*=0.0051), educational attainment (EA) (*p*=8e-10), and intelligence (*p*=7e-08). To assess the association between genetic effects and changes in Swanson, Nolan, and Pelham ADHD Rating Scale-version IV (SNAP-IV) scores, linear mixed-effects models tested the interaction between each PGS and time (baseline, 3-months). PGS for ADHD was associated with higher SNAP-IV scores overall (β=0.1419, p=0.0078, FDR=0.0293) but not with a change at follow-up; whereas, PGS for EA was associated with lower overall scores (β=-0.1161, p=0.0256, FDR=0.0679) and smaller reductions at follow-up (β=0.0946, p=0.0453, FDR=0.1130). Subgroup analyses indicated that these effects were more prominent in adolescents (≥13 years) and individuals classified as responders to treatment. We further explored machine learning models combining clinical and genetic information across 14 models with 49 features. Although models could not empirically generalize, estimated information-theoretic upper bounds for classification accuracy were 72% with PGS, 84% with clinical and demographic variables, and 92% when combined. This study highlights the potential of using PGS and combining genetic and clinical data to improve treatment stratification in ADHD.

## Introduction

Attention deficit hyperactivity disorder (ADHD) is a neurodevelopmental condition with a prevalence estimate of 7.2% among children and adolescents[1], although the likelihood of being diagnosed has increased in recent years. There is compelling evidence of a substantial heritability in the etiology of ADHD, with both rare[2–4] and common[5–7] genetic variants playing a role in the etiology of the condition.

First-line pharmacological treatment for ADHD consists primarily of stimulant medications, which have demonstrated robust efficacy in randomized controlled trials[8, 9]. Methylphenidate is recommended as the initial treatment option in children and adolescents due to its favorable efficacy and tolerability profile[8, 9]. Untreated or inadequately treated ADHD is linked to multiple unfavorable long-term life outcomes, including substance use, antisocial behavior, academic underachievement, and broader psychosocial impairment[10]. In addition to pharmacotherapy, management typically includes interventions such as psychoeducation and environmental adaptations[11].

In our previous prospective clinical study, we found no biological, clinical, or cognitive factors that predicted pharmacological treatment outcomes in children and adolescents with ADHD, highlighting the persistent lack of robust treatment predictors[12]. Genetic factors may partly account for this variability, influencing both symptom presentation and medication response. This predictive difficulty is further complicated by the heterogeneous nature of ADHD[13].

Polygenic scores (PGS) estimate an individual’s common genetic liability to a phenotype by aggregating genome-wide genetic variants weighted by their effect sizes derived from genome-wide association studies (GWAS)[14]. PGSs have been broadly used to investigate genetic liability for psychiatric disorders, including ADHD[15]. More recently, PGS have been investigated as predictors of pharmacological treatment outcomes in ADHD, although findings remain modest and outcome definitions are heterogeneous[16]. In these studies, the treatment outcomes have been measured using clinician-rated global improvement scales with responder classification, short-term symptom change, and register-based measures such as medication discontinuation. In a large Danish register-based study, higher PGS for schizophrenia, major depressive disorder, bipolar disorder, and general psychopathology were associated with increased odds of stimulant discontinuation, whereas higher PGS for cognitive traits – including educational attainment (EA) and intelligence quotient (IQ) – and for body mass index show inverse or age-dependent effects; however, individual PGS explain less than 0.3% of the variance[17]. In 241 Han Chinese children and adolescents with ADHD, PGS for ADHD predicted improvement in ADHD rating scale (ADHD-RS), after 12 weeks of pharmacological treatment, explaining 2.28% of the variance in treatment response; the association was significant for methylphenidate but not atomoxetine[18]. Furthermore, a recent meta-analysis of clinically assessed methylphenidate response as measured by the Global Clinical Improvement Scale across 1000 individuals showed no significant associations with PGS of ADHD or related psychiatric and cognitive traits[19].

To address this complexity, machine learning (ML) approaches are increasingly being applied in ADHD research, as they enable the integration of multidimensional datasets that encompass clinical, genetic, and environmental information. One previous study reported high predictive accuracy (84% using a support vector classifier) for response to methylphenidate by combining traditional clinical measures with genetic polymorphisms located in *ADRA2A* and *SLC6A2* genes[20], using a relatively low sample size (N=78).

Here, we built on our Swedish prospective clinical study[12], by investigating the effects of PGS for ADHD, autism, EA, intelligence and posttraumatic stress disorder (PTSD) on pharmacological treatment response as measured by the change in the Swanson, Nolan, and Pelham ADHD Rating Scale-version IV (SNAP-IV) scale[21] in children and adolescents with ADHD starting ADHD medication. Furthermore, we applied multiple machine learning models integrating phenotypic and genetic predictors and estimated the theoretical upper bound of predictive performance using information-theoretic approaches.

## Materials and methods

### Study cohorts

The primary sample comprised 638 Swedish children and adolescents (6-17 years old) with a clinical ADHD diagnosis initiating medication treatment, recruited through the ADHD medication and predictors of treatment outcome (ADAPT) study (ClinicalTrials.gov Identifier: NCT02136147). Clinical assessments were obtained at baseline and after three months. Detailed cohort characteristics and eligibility criteria have been reported previously by Lilja MM et al[12] and are provided in the Supplementary Methods.

Reference cohorts consisted of 480 and 300 individuals from the Swedish twin studies Gut-2-Twin[22] and the Babytwins Study Sweden (BATSS)[23], respectively. Genotyping, quality control (QC), and PGS calculation were performed using harmonized procedures across cohorts, as detailed below.

All studies were approved by the relevant ethics committees (see in Ethical Information section), conducted in accordance with the Declaration of Helsinki, and written informed consent was obtained from parents or legal guardians.

### Measures

At each visit, parents completed the following questionnaires: the SNAP-IV[21], the Autism Spectrum Screening Questionnaire (ASSQ)[24], the Spence Children’s Anxiety Scale (SCAS)[25], and Pediatric Side Effects Checklist (P-SEC)[26]. IQ was categorized into four groups: above average (≥108), average (92-107), below average (<91), and difficult to assess[27].

The outcome was treatment response, defined as the percentage reduction in SNAP-IV scores after three months relative to baseline[12], classifying participants as responders (≥40%), intermediate responders (20-39%), or non-responders (<20%).

### Polygenic score calculation

Among ADAPT participants, 328 provided saliva samples; and 780 samples were obtained from the reference cohorts Gut-2-Twin and BATSS. DNA was subsequently extracted at the Karolinska Institutet Biobank with the Hamilton ChemagicSTAR® platform.

Genotyping for the three cohorts was performed using the Illumina Infinium assay on the BeadChip GSAMD-24v3-0-EA_20034606_A1, built using the reference genome GRCh37/hg19. The QC for the genotyped data (named target data) was performed using PLINK1.9[28, 29] following the standard procedures implemented in GWAS and PGS studies[30].

Genetic ancestry was estimated using principal component analysis (PCA) based on HapMap III reference data following standard procedures[31, 32], clustering in the PCA was driven primarily by European individuals (Figure S1). Consistently, the linear support vector machine (SVM) classified 300 individuals as European and 9 as non-European. Also, the first four principal components (PCs) values were added in the statistical model to adjust for ancestry. The Haplotype Reference Consortium (HRC), predominantly of European ancestry[33], was used as the reference panel for genotype imputation.

Finally, PGS for ADHD[6], autism[34], educational attainment (EA)[35], intelligence[36] and posttraumatic stress disorder (PTSD) were calculated using GWAS summary statistics as reference data (downloaded in December 2024; Table S1) for the primary analyses. These PGSs were chosen based on prior evidence of associations with ADHD medication treatment response and clinical overlap of the conditions/traits. Furthermore, another 11 GWAS were used for the machine learning models and information-theoretic analysis (Table S1).

An entire description of the sample collection, DNA extraction, genotyping, QC, imputation, and PGS calculation is reported in Supplementary Methods and Table S2.

### Statistical analyses

Group differences in demographic and clinical characteristics were assessed using standard parametric or non-parametric tests, as appropriate. Associations between continuous variables were evaluated using linear regression.

For each of the five PGS (for ADHD, autism, EA, intelligence and PTSD traits), associations with standardized SNAP-IV scores over time were analyzed using linear mixed-effects models (LMM). Models included the interaction between time (baseline versus 3 months) and PGS, and were adjusted for age at study start, sex, and the first four genetic PCs. Random intercepts were included for participant and region. P-values were corrected for multiple testing using the false discovery rate (FDR). Analyses were repeated within age-defined (children: < 13 years old, and adolescents: ≥ 13) and treatment-response subgroups (responders, intermediate responders, and non-responders. Additionally statistical details are provided in Supplementary Methods.

### Machine learning and information-theoretic analysis

Predictors consisted of phenotypic characteristics and 16 PGS (Table S3). The outcome variable was treatment response, categorized as responder, intermediate responder, or non-responder. Data were stratified into training, validation, and test sets, and preprocessing was performed using parameters estimated from the training data.

We evaluated multiple machine learning algorithms, including logistic regression, decision trees, random forests, support vector machines, k-nearest neighbors, naïve Bayes classifiers, perceptron, and XGBoost[37]. Predictors were ranked according to their mutual information, and progressively larger feature subsets were constructed from the highest-ranked predictors.

To estimate the information-theoretic upper bound on performance, we applied Fano’s inequality. Joint mutual information between predictors and outcome was approximated using the tensorization upper bound based on the sum of individual feature mutual information values[38]. Features were grouped into genetic (PGS) and non-genetic (clinical, demographic, and phenotypic) categories, and combined sets.

Model performance was optimized on the validation set and best model was subsequently evaluated on an independent test set. Statistical significance was assessed using a label permutation test with 100 random permutations. Additional methodological details are provided in the Supplementary Methods.

## Results

### Sample description

We analyzed a subset of 328 children and adolescents from the ADAPT cohort based on availability of genotyping and clinical data for baseline and 3-months follow-up. After QC, 309 individuals were included in the statistical analysis. Individuals with genotyped samples that passed the QC were compared with the remaining participants in the cohort (Table 1, Table S4); differences were observed only in the region where they came from (*p*-value=2.25e-10).

**Table 1.** Group differences in the main clinical variables used in this study between individuals with genotyped samples that passed the QC and the remaining participants in the cohort.

| Variable | Genotyped <sup>a</sup> | Not genotyped <sup>a</sup> | *p*-value |
| --- | --- | --- | --- |
| Age start | 11.55±0.29 | 11.44±0.18 | 0.738 |
| Sex (Female) | 109 (35.3%) | 42 (38.2%) | 0.667 |
| SNAP-IV total (baseline) | 47.17±1.88 | 49.97±1.02 | 0.193 |
| SNAP-IV total (month 3) | 34.85±1.67 | 35.46±1.01 | 0.754 |
| ASSQ total (baseline) | 10.68±0.75 | 11.11±0.46 | 0.627 |
| ASSQ total (month 3) | 8.86±0.74 | 9.03±0.43 | 0.842 |
| **<u>Outcome</u>** |  |  | 0.787 |
| • Non-responder | 118 (38.2%) | 46 (41.8%) |  |
| • Intermediate responder | 89 (28.8%) | 29 (26.4%) |  |
| • Responder | 102 (33%) | 35 (31.8%) |  |
| **<u>Regions</u>** |  |  | 2.57E-09 |
| • Gotland | 16 (5.2%) | 2 (1.8%) |  |
| • Stockholm | 207 (67%) | 104 (94.5%) |  |
| • Västerbotten | 86 (27.8%) | 4 (3.6%) |  |
| **<u>IQ collapsed (baseline)</u>** |  |  | 0.500 |
| • Below average (IQ = 69-92) | 61 (35.3%) | 16 (28.6%) |  |
| • Average (IQ = 93-107) | 74 (42.8%) | 23 (41.1%) |  |
| • Above average (IQ = 108-131) | 28 (16.2%) | 14 (25%) |  |
| • Difficult to assess | 10 (5.8%) | 3 (5.4%) |  |
| **<u>Medication</u>** |  |  | 0.767 |
| • Atomoxetine | 16 (7.2%) | 5 (7.9%) |  |
| • Combined methylphenidate & atomoxetine | 1 (0.5%) | 1 (1.6%) |  |
| • Guanfacine | 3 (1.4%) | 0 (0%) |  |
| • Lisdexamfetamine | 43 (19.5%) | 12 (19%) |  |
| • Methylphenidate | 158 (71.5%) | 45 (71.4%) |  |

<sup>a</sup> For continuous variables, values are presented as mean ± standard error of the mean (SEM). For categorical variables, data are presented as counts and percentages.

First, we compared the five calculated PGSs of the ADAPT cohort with the scores of a reference sample derived from two Swedish population-based twin cohorts to test whether their genetic liability for these traits differ from a population mean (Figure 1A, Figure S2A-S2B). We show that the ADAPT cohort had a significantly higher PGS for ADHD compared with the reference cohorts (*p*<2.22e-16); but significantly lower PGS for autism (*p*=0.0035), EA (*p*=5.2e-09), and intelligence (*p*=1.4e-08). As the results for PGS for autism were surprising, we separated the reference cohort to the two samples showcasing that the ADAPT cohort had lower PGS for autism than the Gut-2-Twin reference cohort (*p*<2.22e-16, N=480) but higher PGS than the BATSS reference cohort (*p*=7.4e-08, N=300).

**Figure 1.**
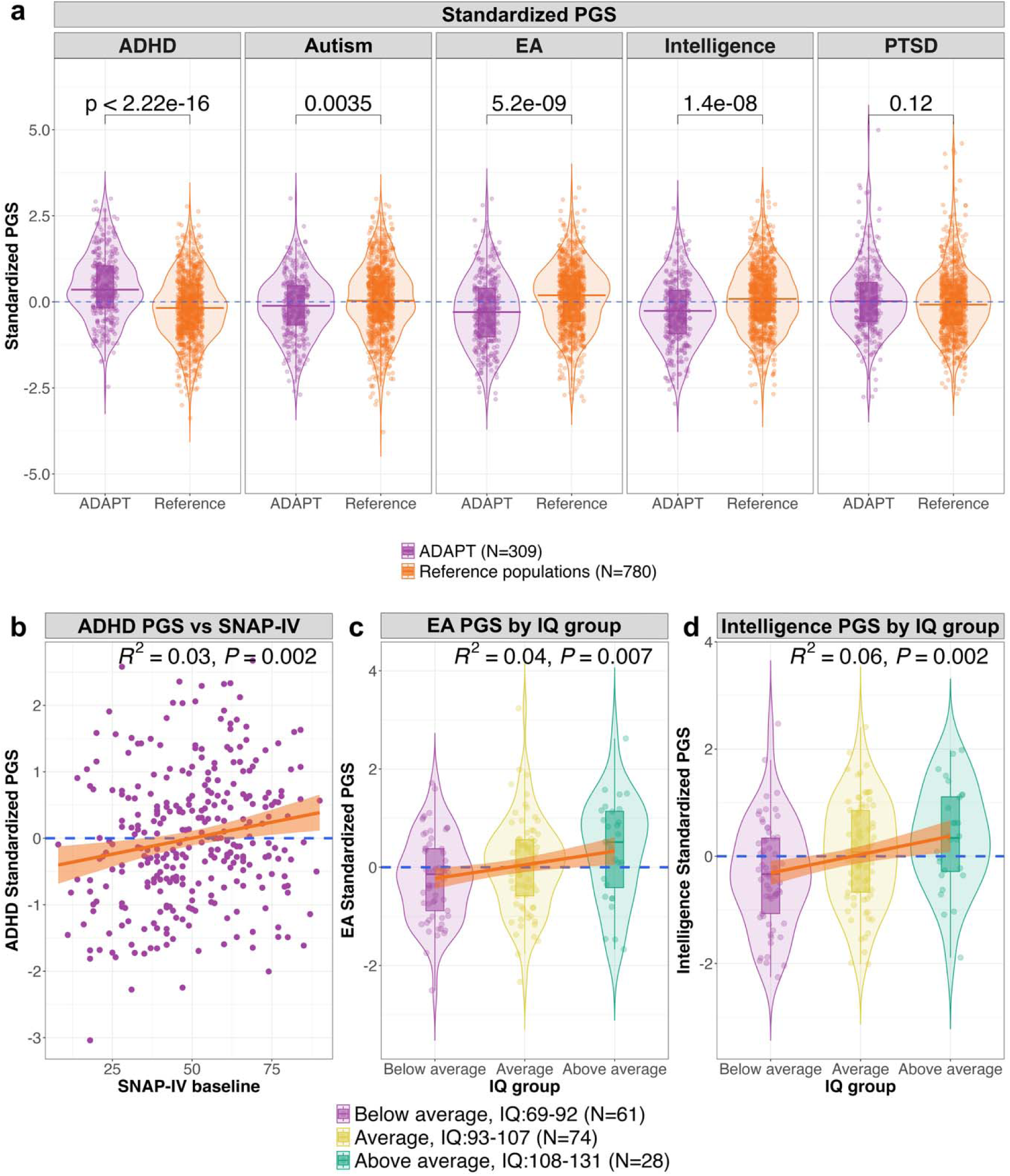
Polygenic scores (PGS) across cohorts and clinical correlations at baseline. **(A)** Comparison between ADHD, autism, EA, intelligence, and PTSD PGS in ADAPT (N=309) and reference cohorts (Gut-2-Twin and BATSS, N=780); p-values from Student’s t-test are shown. **(B)** Association between PGS for ADHD and the SNAP-IV score in ADAPT. **(C-D)** Associations of PGS for EA and intelligence with IQ in ADAPT (N=163 with available data). Orange lines represent the fitted linear models with 95% confidence interval

### Associations between PGSs and clinical features

Associations between PGS and related clinical phenotypes at baseline were assessed separately for the five genetic traits studied (ADHD, autism, EA, intelligence, and PTSD) to evaluate whether the calculated PGS captured the expected trait-specific genetic liability. We demonstrate a positive significant correlation between PGS for ADHD and the SNAP-IV (R^2^=0.03, *p*=0.002) (Figure 1B). Furthermore, both PGS for EA and intelligence had positive associations with the IQ level groups (R^2^=0.04, *p*=0.007; and R^2^=0.06, *p*=0.002; respectively; Figure 1C-D). However, we did not find significant correlations between the PGS for autism and ASSQ scores, or between the PGS for autism and the ASSQ cut-off for autism (Figure S2C-D).

Additionally, we correlated the different PGS between them (Figure S3). PGS for ADHD had a positive correlation with PGS for autism (R^2^=0.11, FDR *p*=4.4e-09) and for PTSD (R^2^=0.21, FDR *p=*8.4e-17). As expected, we observe a negative correlation between PGS for ADHD and PGS for EA (R^2^=0.10, FDR *p*=2.1e-08). Finally, no correlation was observed between PGS for ADHD and PGS for intelligence (R^2^=0, FDR *p*=0.43).

### Associations between PGSs and reductions in SNAP-IV over time

To determine whether genetic liability is associated with changes in the SNAP-IV scale as a measure for ADHD treatment outcome, we fitted LMM, testing the interaction between each PGS and time (baseline and 3-months follow-up). Among the five PGSs tested (Figure 2A, Table S5), PGS for ADHD showed a significant association with higher SNAP-IV values at both time points (β=0.1419 [0.0378–0.2461], *p*=0.0078, FDR *p*=0.0293) but no association with the change. None of the other tested PGSs had a significant association with SNAP-IV values or with the change after FDR p-value corrections. Nominal association was found for the PGS for EA, which was associated with lower SNAP-IV values overall (β=-0.1161 [-0.2178–-0.0145], *p*=0.0256, FDR *p*=0.0679) as well as with lower SNAP-IV reduction over the 3-months follow-up (β=0.0946 [0.0024–0.1868], *p*=0.0453, FDR *p*=0.1133). In all the models, a significant association was observed between SNAP-IV scores and principal component 4 (PC4; accounting for 2.61% of the explained variance), reflecting ancestry-related effects.

**Figure 2.**
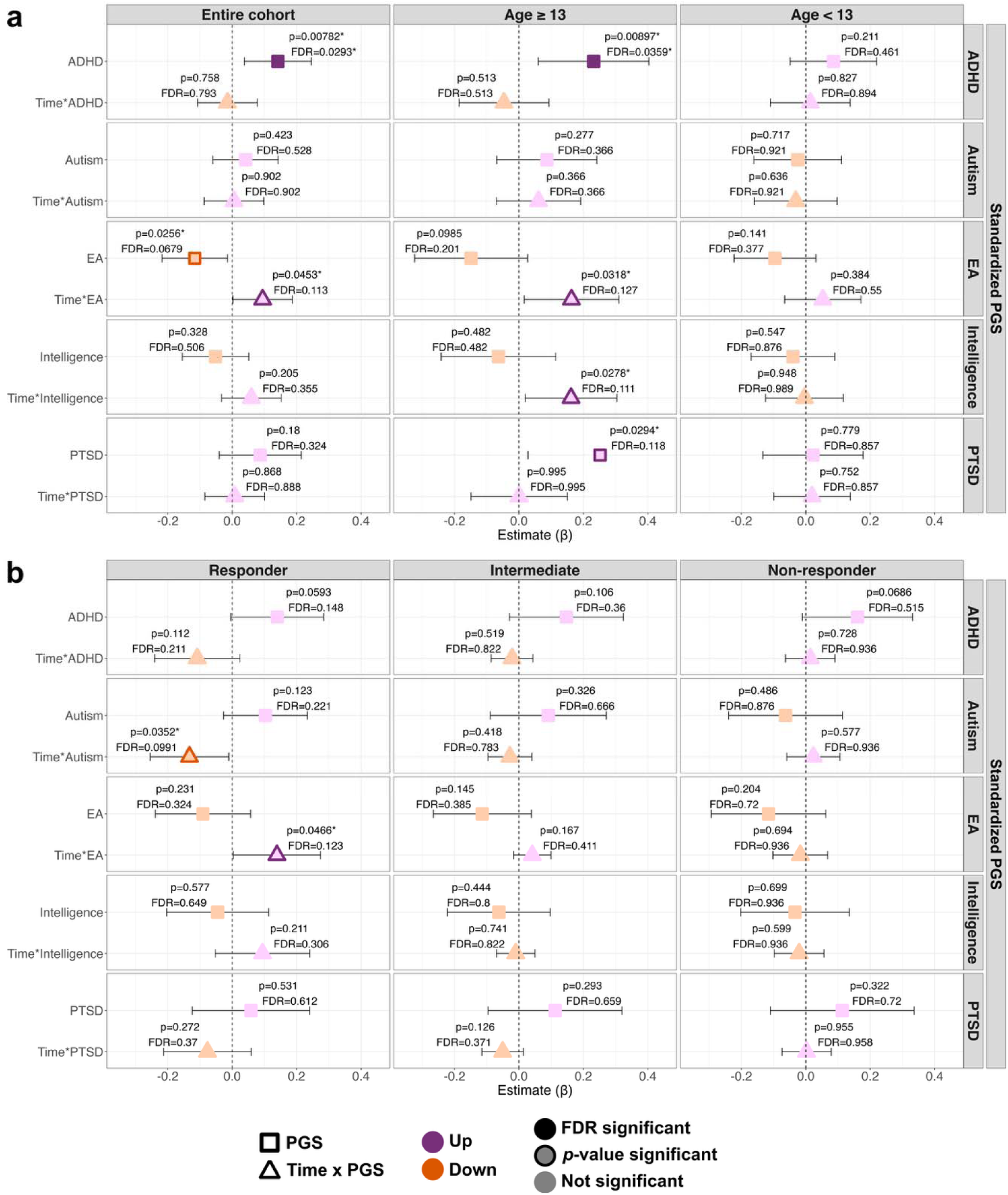
Associations between polygenic scores (PGS) and SNAP-IV scores in the ADAPT cohort (N=309). Linear mixed-effects models tested associations between standardized PGS and SNAP-IV scores over time (baseline vs 3 months). **(A)** The entire cohort and stratified by age (≥13 and <13 years). **(B)** By treatment outcome (responders, intermediate responders, and non-responders). Points show estimates (β) with 95% confidence intervals. Shapes and colors indicate model term, effect direction, and significance

To investigate potential age effects, we fitted separate LMMs for adolescents (≥13 years old) and children (<13), considering the significant correlation between age and SNAP-IV at baseline (R^2^=0.07, FDR *p<*0.001). No significant associations were observed in the younger group (Figure 2A, Table S5). In the older group, we replicated the findings from the full cohort: higher PGS for ADHD predicted higher SNAP-IV values (β=0.2319 [0.0601–0.4037], *p*=0.0090, FDR *p*=0.0359), and higher PGS for EA had non-significant association with reduced SNAP-IV change over time (β=0.1630 [0.0159–0.3101], *p*=0.0318, FDR *p*=0.1272). Moreover, the association between higher PGS for intelligence and lower SNAP reductions did not hold for FDR correction (β=0.1617 [0.0194–0.3041], *p*=0.0278, FDR *p*=0.1113). Furthermore, in the older group, we saw a higher PGS for PTSD with greater SNAP-IV values, but it did not survive the multiple correction (β=0.2520 [0.0275–0.4766], *p*=0.0294, FDR *p*=0.1177).

Lastly, we stratified by the outcome-defined subgroups and fitted three separate LMMs for each group, using the same model framework. No significant associations were identified in the intermediate or non-responder groups (Figure 2B, Table S6). Among responders, higher PGS for EA had non-significant association with lower SNAP-IV reduction over time (β=0.1391 [0.0038–0.2744], *p*=0.0466, FDR *p*=0.1235), in line with the findings in the entire cohort and in the older individuals, whereas higher PGS for autism were associated with greater improvement between baseline and follow-up (β=-0.1325 [-0.2542–-0.0108], *p*=0.0352, FDR *p*=0.0991). These associations did not withstand FDR correction. Additionally, we stratified the individuals into tertiles based on their PGS for ADHD and EA to evaluate associations with genetic liability and treatment response. Higher PGS for ADHD was associated with higher SNAP-IV scores, whereas higher PGS for EA was associated with lower SNAP-IV scores (Figure S4). However, the reduction in SNAP-IV scores from baseline to 3-month follow-up was comparable across PGS for ADHD tertiles. In contrast, individuals in the lowest PGS for EA tertile exhibited a greater reduction in SNAP-IV scores than those in the higher tertiles (Figure S4).

Across the entire cohort and stratified by age models, PGS and covariates explained a modest proportion of variance in SNAP-IV scores through time (marginal R^2^=0.1965±0.0134 [mean±SD]; Figure S5). Stratified analyses by treatment outcome revealed differences in the marginal explanatory power (responders=0.5697±0.0029, intermediate responders=0.3089±0.0055, non-responders=0.0800±0.0107; Figure S5).

### Integrating genetic and non-genetic data enhances theoretical prediction of medication response

Even though our selected PGS did not reach FDR-corrected associations, several PGS may contribute with minor effects to the treatment outcomes. We, therefore, fitted 14 models with 49 genetic and non-genetic features to assess whether their combination provided additional information for predicting the three groups by treatment outcome.

While many features contributed little mutual information, some predictors contributed substantially more than others (Figure 3A), resulting in higher theoretical accuracy bounds (Figure 3B). Using only PGS values, the estimated information-theoretic upper bound on classification accuracy was 72%. Using only non-genetic clinical and demographic variables, the corresponding upper bound increased to 83%. Combining genetic and non-genetic predictors further increased the estimated upper bound to 93%.

**Figure 3.**
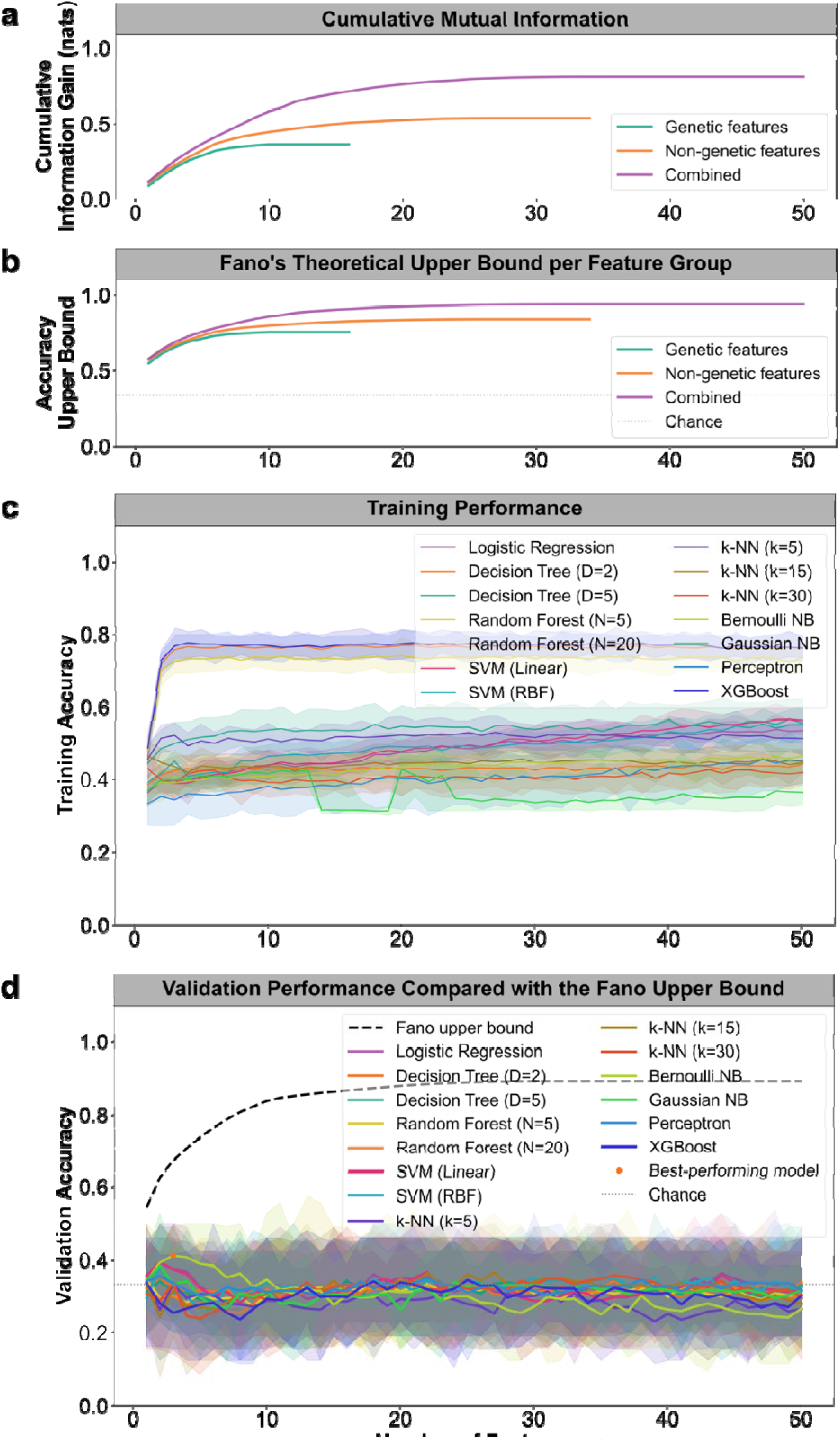
Theoretical and observed prediction of medication response. **(A)** Cumulative mutual information for genetic, non-genetic, and combined feature sets. **(B)** Information-theoretic upper bounds on classification accuracy as a function of the number of features. **(C)** Training performance of machine learning models across increasingly large feature sets. **(D)** Validation performance and confidence intervals

These model-independent theoretical limits were not approached in practice. Although several models achieved varying levels of training performance (Figure 3C), validation performance remained substantially lower (Figure 3D), indicating limited generalization from the available data. Similarly, the final model achieved a test accuracy of 24%. This is lower than the accuracy obtained under the random label permutation test (34%), and especially the 95^th^ percentile of the permutation distribution (48%). Consequently, we could not reject the null hypothesis that there is no association between model predictions and the true treatment-response labels.

## Discussion

This prospective cohort provides an opportunity to investigate the contribution of genetic factors to pharmacological treatment response in pediatric ADHD, using clinical data collected before and after treatment initiation, alongside genetic data. As expected, our cohort of children and adolescents with ADHD showed higher PGS for ADHD than reference cohorts. Within the ADHD cohort, individuals with higher PGS for ADHD showed higher SNAP-IV scores, whereas individuals with higher PGS for EA exhibited lower SNAP-IV scores. Higher PGS for EA was also nominally associated with reduced symptom improvement over time.

Our results demonstrated a significant correlation between PGS for ADHD and baseline SNAP-IV scores in children and adolescents with ADHD, even though the effect size was small. Higher PGS for ADHD was associated with more severe baseline symptoms, particularly in adolescents (≥13 years old). This age-specific effect may reflect greater stability of ADHD symptoms as patients transition into adolescence, or that children’s symptoms are more dependent on environmental factors[39, 40].

The lower PGS for autism in ADHD cohort compared with reference cohorts was unexpected given the high comorbidity between ADHD and autism[41–43], although the effect size of this difference is smaller than the other expected effects. This finding could arise from enrollment bias in the reference cohorts, as participation is voluntary. Notably, Gut-2-Twin showed higher PGS for autism than ADAPT and BATSS lower, suggesting cohort-specific differences in autism genetic liability. The absence of a significant correlation between PGS for autism and ASSQ further complicates the interpretation of this finding, which may reflect differences in phenotype definition, as the autism GWAS was based on clinical diagnosis[34], and ASSQ measures the severity of autistic symptoms[24]. Consistent with previous findings from this cohort, PGS for autism was not associated with treatment outcome in the entire cohort[12, 44]. Although co-occurring autism has been proposed to reduce ADHD medication effectiveness[45], individuals with ADHD and autism are less likely to initiate pharmacological treatment[46], experience longer delays to treatment initiation, and show higher rates of treatment discontinuation than those with ADHD alone[47]. Against this background, we observed a nominal association between higher PGS for autism and greater symptom improvement among responders.

The lower PGS for EA and intelligence in ADAPT than in the reference cohorts are consistent with previous evidence linking ADHD genetic liability to poorer educational and cognitive outcomes[48, 49]. Whereas a previous study in an independent ADHD cohort reported positive associations between IQ and methylphenidate response[50–52]; neither IQ in previous analysis of ADAPT cohort[12] nor PGS for intelligence in the present study was associated with symptom improvement. In contrast, higher PGS for EA was nominally associated with smaller reductions in SNAP-IV over time, indicating less symptom reduction. This observation is noteworthy given previous evidence that PGS for EA captures genetic and phenotypic heterogeneity within ADHD[53], and may influence treatment-related outcomes[17]. One possible explanation is that patients with high PGS for EA present ADHD manifestations that are less well captured by SNAP-IV, consistent with the observed negative correlation between PGS for EA and baseline symptom severity. However, the finding could also suggest reduced responsiveness to psychopharmacological treatment. Interestingly, the opposite effect have been seen for other types of interventions as reports have linked higher PGS for EA to greater reductions in anxiety and depressive symptoms, including internet-delivered cognitive behavior therapy[54], antidepressant treatment[55], and schizophrenia rehabilitation[56].

Being able to predict treatment response before medication initiation would be valuable for both clinical practice and research. Although several clinical variables and PGSs had nominal associations with treatment outcome, prediction performance remains poor. The information-theoretic analysis suggested that limited predicted accuracy was not solely explained by the information content of the available predictors, highlighting potential contributions of sample size, feature representation, and model limitations. Nevertheless, combining genetic and non-genetic variables modestly increased the estimated predictive potential, supporting the value of integrating both sources of information in future prediction models.

In conclusion, this study highlights the complexity of predicting pharmacological treatment response in ADHD. In this study, we corroborated the PGS data to validate the ADAPT cohort as having a specific genetic liability for ADHD. We observed associations between the PGS for EA and both symptom severity and treatment response, with a stronger effect on treatment response, particularly among adolescents, where higher genetic liability to EA was nominal associated with poorer outcomes. We found no evidence of associations between PGSs for autism, IQ, PTSD, or ADHD and treatment response. Importantly, integrating genetic information with clinical variables modestly improved the estimated information-theoretic upper bound on prediction performance of treatment response, supporting the potential value of machine learning models using not only traditional clinical features but also PGS data. Future research should focus on integrating PGS with environmental factors and individual-level clinical data to improve the identification of patients most likely to benefit from ADHD medication. Treatment responders represent the most clinically relevant subgroup and appear to be more homogenous, supporting the potential value of further characterizing responders to improve stratification and treatment selection in ADHD. Finally, future studies with larger sample sizes and more powerful GWAS will be essential for refining these predictions and improving their clinical applicability.

## Data Availability

The analyses presented here were preregistered (ClinicalTrials.gov Identifier: NCT02136147). The data presented here are not publicly accessible but will be made available upon reasonable request to the corresponding author. Note that sharing of pseudonymized personal data will require a data sharing agreement, according to Swedish law. The code necessary to reproduce the analyses from this study is available in GitHub (https://github.com/Tammimies-Lab/ADAPT_ADHD_PGS_2026).

## Author contributions

**Alba Escalera-Balsera:** Conceptualization, Methodology, Data curation, Formal analysis, Visualization, Writing – original draft, Reviewing and Editing.

**Maria M Lilja:** Conceptualization, Methodology, Data curation, Writing, Reviewing and Editing.

**Mattias Nordstrand:** Methodology, Software, Investigation, Data curation, Formal analysis, Visualization, Writing – original draft, Review and Editing.

**Terje Falck-Ytter:** Data curation, Writing, Reviewing and Editing.

**Eva Serlachius:** Conceptualization, Reviewing and Editing.

**Jyoti Bhagia:** Conceptualization, Methodology, Reviewing and Editing.

**Kristiina Tammimies:** Conceptualization, Methodology, Funding acquisition, Project administration, Writing, Reviewing and Editing.

**Linda Halldner:** Conceptualization, Methodology, Funding acquisition, Project administration, Writing, Reviewing and Editing.

## Ethical information

Written informed consent was obtained from all caregivers, and the research was conducted in accordance with the Declaration of Helsinki. The Regional Ethical Board in Stockholm (Dnr 2013/1007-31, 2016/1350-32, 2018/2404) and the Swedish Ethical Review Authority (Dnr 2021-06658-02) approved the ADAPT study, the BATSS study (2014/2248-32) and the Gut-2-Twin study (2020-03226).

