## Supplementary material for "Polygenic and machine learning analysis of medication response in a prospective pediatric cohort initiating ADHD medication"

### Supplementary information

### Supplementary Methods

#### Study individuals

Participants from the ADHD medication and predictors of treatment outcome (ADAPT) study (ClinicalTrials.gov Identifier: NCT02136147), were recruited and the data collected from July 2014 to June 2022 from 15 Swedish Child and Adolescent Psychiatry outpatient units in three different regions in Sweden: Stockholm (13 units), Gotland (one), and Västerbotten (one).

ADAPT was designed as a prospective observational cohort study, where all the individuals were about to initiate ADHD medication (methylphenidate, dexamfetamine, lisdexamfetamine, atomoxetine, or guanfacine). None of the participants had received ADHD medication within the three months prior to study inclusion.

Written informed consent from parents or legal guardians were collected. The study was performed in line with the principles of the Declaration of Helsinki. The ADAPT study was approved by the Regional Ethical Board in Stockholm (Dnr 2013/1007-31, 2016/1350-32, 2018/2404), and by the Swedish Ethical Review Authority (Dnr 2021-06658-02).

#### Measures

At each visit, parents completed the following questionnaires: the SNAP-IV[24], the Autism Spectrum Screening Questionnaire (ASSQ)[25], the Spence Children's Anxiety Scale (SCAS)[26], and Pediatric Side Effects Checklist (P-SEC)[27]. Additionally, information on the IQ level of each participant was retrieved and consolidated into four groups: above average (IQ ≥ 108), average (92 ≤ IQ < 107), below average (IQ < 91) and difficult to assess[28], which was studied together with the missing values.

In the previous paper, the treatment effect was defined as the reduction in ADHD symptoms, as measured by the SNAP-IV scale after three months of treatment, relative to baseline[15]. Based on this definition, participants were divided into three groups by outcomes defined as responders (SNAP-IV reduction ≥ 40%), intermediate responders (20% ≤ SNAP-IV reduction < 40%) and non-responders (SNAP-IV reduction < 20%).

#### Sample collection and DNA extraction

In the ADAPT cohort, 328 saliva samples were collected using the Oragene DNA OG-500 collection tubes (DNA Genotek, Inc., Ottawa, Ontario, Canada). In the reference cohorts Gut-2-Twin and BATSS, 780 saliva samples were collected using the DNA Genotek OG-575 collection kit as described earlier. DNA was subsequently extracted at the Karolinska Institutet Biobank with the Hamilton ChemagicSTAR® platform. DNA quantity and quality were assessed before genotyping.

Because the reference cohorts consisted of twin pairs, one individual from each pair was selected for the analysis. Sample identifiers were encoded using the same alphanumeric code for each pair twin, followed by either "1" or "2" to denote the individual twin. Therefore, the twin labeled "1" was selected whenever both individuals from a twin pair had passed the quality control (QC). In cases where only the twin labeled "2" has passed QC, that individual was selected instead.

### Genotyping and quality control

Genotyping was performed using the Illumina Infinium assay on the BeadChip GSAMD-24v3-0-EA_20034606_A1, which contains 730,059 SNP probes. The results were analyzed using the software GenomeStudio 2.0.3. It was built using the reference genome GRCh37/hg19, and the positions of each SNP were determined from the corresponding manifest for this BeadChip. The QC for the genotyped data (named target data) was performed using PLINK1.9[1, 2] following the standard procedures implemented in GWAS and PGS studies[3]. Both single-nucleotide polymorphisms (SNPs) and individuals were first filtered for missingness > 20%, followed by a second filtering step at > 2%. Sex discrepancies were then assessed in two cases, PLINK1.9 classified individuals as "unknown", and the reported sex was therefore retained. SNPs located on sex chromosomes were removed. Only common variants were retained by filtering for minor allele frequency (MAF) ≥ 0.05. To ensure adherence to the Hardy-Weinberg equilibrium, variants with p-values < $1x10^{-6}$ were filtered out. SNPs were pruned for linkage disequilibrium (LD) using an $r^2$ threshold > 0.2, and individuals with excessive heterozygosity were removed by F coefficient passing ± standard deviations (SD) from the mean. Finally, duplicated variants were excluded. Genotyping of the reference cohorts was performed using the same pipeline as that used for the ADAPT cohort. After QC, 93,237 variants were retained in the ADAPT cohort, 92,332 in Gut-2-Twin, and 93,980 in BATSS.

Genetic ancestry was estimated using principal component analysis (PCA) based on HapMap III reference data, following standard procedures[4, 5]. Prior to PCA, the reference dataset underwent QC, including the exclusion of ambiguous SNPs (non-A/T or non-C/G), the retention of variants shared with the target dataset, and the correction of strand mismatches. European ancestry was predicted using a linear support vector machine (SVM) trained on the first four principal components (PCs) of genetic variation (83.54% explained). The model was trained on a reference dataset (80%) and evaluated on a test set (20%), achieving 100% classification accuracy. The trained model was subsequently applied to classify ancestry in the ADAPT cohort. Moreover, the first four PCs values were added in the statistical model to adjust for ancestry. Parameters and thresholds applied during the QC are reported in Table S2.

### Imputation

The Haplotype Reference Consortium (HRC), predominantly of European ancestry[6], was used as the reference panel for genotype imputation. Before imputation, chromosomes were harmonized to ensure consistent sample sets across all chromosomes, and sex chromosomes were excluded.

For the target dataset, pre-phasing, phasing, and imputation steps were performed using imputeInversion[7], SHAPEIT5[8] and IMPUTE5[9], respectively. Finally, variant annotation was performed to assign the corresponding rsID using the ANNOVAR hg19 dataset[10]. Imputation of the reference cohorts was carried out using the same pipeline as for the ADAPT cohort. Genotype imputation yielded a total of 39,131,578 variants.

Post-imputation QC was subsequently performed using the same filtering criteria for missingness, MAF, and the Hardy-Weinberg equilibrium described above; resulting in 5,294,765 variants retained in the ADAPT cohort for downstream analyses, 5,210,361 in Gut-2-Twin, and 5,243,872 BATSS.

### Polygenic score calculation

PGS for ADHD[11], autism[12], educational attainment (EA)[13], intelligence[14] and posttraumatic stress disorder (PTSD) were calculated using GWAS summary statistics as reference data (downloaded in December 2024; Table S1) for the primary analyses. These PGSs were chosen as there was earlier evidence of ADHD medication treatment response and clinical overlap of the conditions/traits. Furthermore, GWAS for the following traits were used for the machine learning models and information-theoretic analysis (Table S1): ADHD[11], anxiety[15], agreeableness[16], autism[12], circadian rhythms[17], conscientiousness[16], depression[18], extraversion[16], EA[13], insomnia[19], intelligence[14], neuroticism[16], openness[16], PTSD[20], short sleep duration[21], and long sleep duration[21].

To minimize the impact of population stratification, only individuals with European ancestry from the GWAS reference samples were included. As described above, the base reference datasets were filtered to retain common variants (MAF ≥ 0.05), variants with imputation quality INFO ≥ 0.08, and to exclude duplicated and ambiguous SNPs.

For each reference dataset, the natural logarithms of the estimated odds ratios (OR) and the corresponding p-values for each SNP allele were used. PGS were calculated by summing allele dosages (0, 1, 2), weighted by the natural logarithm of the allelic OR, to obtain a genome-wide aggregate effect. The SNP effect size was estimated using PRS-CS[22] (parameters used in Table S2), and the final PGS was calculated using the --score function in PLINK1.9[1, 2]. Finally, PGS were standardized (mean = 0, SD = 1) to perform association testing.

### Statistical analysis

Categorical variables were compared between groups using the chi-square test, with Fisher's exact test applied when expected counts were small. Continuous variables were assessed for normality within each group (Shapiro-Wilk test) and compared using the Student's t-test or Wilcoxon rank-sum test as appropriate. Associations between continuous variables and between continuous outcomes and ordinal or categorical groups (treated numerically), were evaluated using linear regression, reporting the coefficient of determination ($R^2$) and corresponding *p*-values.

For each of the five PGS (for ADHD, autism, EA, intelligence and PTSD traits), associations with standardized SNAP-IV scores over time were analyzed using linear mixed-effects models (LMM). The models included the interaction between time (baseline versus 3 months) and PGS, as well as the main effect of the PGS; they were adjusted for age at study start, sex, and the first four genetic principal components as fixed effects. Random intercepts were included for participants and for the region to account for repeated measurements and potential regional clustering. Model estimates, standard errors (SE), and p-values were extracted for all fixed effects; p-values were corrected for multiple testing using the false discovery rate (FDR). These analyses were also repeated within subgroups, using the same LMM structure applied. First, participants were separated by age into adolescents (≥ 13 years old) and children (< 13 years old); in these models, all covariates were included except age. Second, participants were divided by treatment outcome into responders, intermediate responders, and non-responders, as explained previously.

All statistical analyses were performed using R (v4.4.2), using *ggplot2*[23], *ggpubr*[24], *dplyr*[25] and *tidyr*[26] packages for the visualizations.

### Machine learning and information-theoretic analysis

Predictors consisted of phenotypic characteristics and 16 PGS (Table S3). The outcome variable was treatment response, categorized as responder, intermediate responder, or non-responder, similar to the main analyses. Participants considered likely to have received medication despite missing medication status were included only in the training set to increase training data while preserving reliable validation and test evaluation.

The remaining data were stratified into training, validation, and test sets, with approximately 10% reserved for testing and the remainder split into training and validation subsets (90%/10%). Missing numerical and categorical values were imputed using the training set median and the most frequent category, respectively. Numerical variables were standardized using a StandardScaler fitted on the training data, while categorical variables were encoded using an OrdinalEncoder.

Analyses were conducted in Python 3.12.3 using scikit-learn 1.7.0[27], NumPy 2.0.0[28], and pandas 2.2.3[29, 30]. Machine learning models included logistic regression, decision trees, random forests, support vector machines, k-nearest neighbors, naïve Bayes classifiers, perceptron, and XGBoost[31].

Mutual information between each predictor and treatment outcome was estimated on the training set. Predictors were ranked by mutual information, and progressively larger feature subsets were constructed from the highest-ranked predictors. Mutual information was calculated for discrete variables empirically, and mutual information for continuous variables was estimated using a k-nearest-neighbor estimator (k = 3)[32–34], as implemented in scikit-learn's mutual_info_classif function. To account for stochastic variation introduced by the estimator, the mutual information estimation was repeated using 100 random seeds. The upper 95$^{th}$ percentile of the resulting mutual information estimates was used as an optimistic estimate of the available information when deriving an information-theoretic upper bound on prediction accuracy. Features were grouped into genetic (PGS) and non-genetic (clinical, demographic, and phenotypic) categories, and cumulative mutual information was evaluated separately for genetic, non-genetic, and combined feature sets.

To estimate the information-theoretic upper bound on performance, we applied Fano's inequality. As direct estimations of the joint mutual information $I(Y;X)$ are infeasible in high-dimensional settings with limited sample sizes, we approximate it using the tensorization[35] upper bound

$$I(Y;X) \leq \sum_{i=1}^{K} I(Y;X_i)$$

which assumes conditional independence among the predictors given the outcome. Since this assumption rarely holds in practice, the estimate represents an optimistic upper bound estimate on the available information. Conditional entropy was estimated as

$$H(Y|X) = H(Y) - I(Y;X)$$

where $H(Y)$was calculated by the class distribution. A bisection search was then used to find the smallest error probability $P_e$ satisfying Fano's inequality

$$H_b(P_e) + P_e \log(|Y| - 1) \geq H(Y|X)$$

where $H_b(\cdot)$ denotes the binary entropy function. The corresponding theoretical upper bound on classification accuracy was estimated as $1 - P_e$.

For each model, validation accuracy was evaluated as a function of the number of top-ranked features. Bootstrap resampling (50 iterations) was used to estimate mean validation performance and 95% confidence intervals using feature subsets of increasing sizes. The best-performing model and feature subset were retrained using the combined training and validation data and evaluated on the independent test set. Statistical significance was assessed using a label permutation test with 100 random permutations of the test labels while keeping the model predictions fixed.

### Code availability

The code necessary to reproduce all the analyses from this study is available in GitHub (https://github.com/Tammimies-Lab/ADAPT_ADHD_PGS_2026).

### Supplementary Tables

***Table S1.*** *Description of the base GWAS studies used for polygenic scores (PGS) calculation.*

| Phenotype | GWAS study | Number of participants | | | Used in main analysis |
|---|---|---|---|---|---|
| | | **Cases** | **Controls** | **Total** | |
| ADHD | Demontis D, et al. 2023 | 38,691 | 186,843 | 225,534 | Yes |
| Anxiety | Friligkou E, et al. 2024 | 87,517 | 1,096,458 | 1,183,975* | No |
| Autism | Grove J, et al. 2019 | 18,381 | 27,969 | 46,350 | Yes |
| Depression | Major Depressive Disorder Working Group of the Psychiatric Genomics Consortium. 2025 | 547,355 | 2,071,918 | 2,619,273** | No |
| EA | Lee JJ, et al. 2018 | 766,345 | | 766,345** | Yes |
| Intelligence | Savage JE, et al. 2018 | 269,867 | | 269,867 | Yes |
| Agreeableness | Gupta P, et al. 2024 | 252,749 | | 252,749 | No |
| Conscientiousness | | 252,253 | | 252,253 | No |
| Extraversion | | 298,772 | | 298,772 | No |
| Neuroticism | | 682,688 | | 682,688 | No |
| Openness | | 237,390 | | 237,390 | No |
| PTSD | Nievergelt CM, et al. 2024 | 150,793 | 1,130,197 | 1,280,990 | Yes |
| Insomnia | Watanabe K, et al. 2022 | 109,548 | 277,440 | 386,988** | No |
| Sleep duration: short (≤ 5h) | Austin-Zimmerman I, et al. 2023 | 18,915 | | 445,966 | No |
| Sleep duration: long (≥ 10h) | | 6,521 | | 445,966 | No |
| Circadian rhythms | Jones SE, et al. 2019 | 252,287 (morning people) | 150,908 (evening people) | 403,195 | No |

*The number of samples reported in this table is the number of samples used in this project analyses.*

**The GWAS include samples from a global ancestry, but for this study we are using only the samples with European ancestry.*

***The GWAS include samples for 23andMe consortium, but it is not possible to use their data without 23andMe approval. Therefore, the samples from 23andMe are excluded in this study.*

*Attention Deficit Hyperactivity Disorder (ADHD), Educational attainment (EA), Posttraumatic stress disorder (PTSD).*

***Table S2.*** *Parameters used in quality control (QC)and polygenic score (PGS) calculation.*

| **Parameter used in Plink for QC** | | |
|---|---|---|
| **Parameter** | **Threshold** | **Description** |
| --keep-allele-order | | Forces the original A1/A2 allele encoding to be preserved. |
| --mind | first 0.2, then 0.02 | Filters out all samples with missing call rates exceeding the provided value to be removed. |
| --geno | first 0.2, then 0.02 | Filters out all variants with missing call rates exceeding the provided value to be removed. |
| --maf | 0.05 | Filters out all variants with minor allele frequency below the provided threshold. |
| --hwe | 1.00E-06 | Filters out all variants which have Hardy-Weinberg equilibrium exact test p-value below the provided threshold. |
| --indep-pairwise | 50 5 0.2 | Keep markers that are in approximate linkage disequilibrium with each other. Parameters: window size [variant count], variant count to shift the window at the end of each step, pairwise $r^2$ threshold. |
| --list-duplicate-vars | suppress-first | Identify duplicated variants, keeping the first variant in each group of duplicates. |
| --het | | Individuals with heterozygosity rates out of the range of ± 3 standard deviations of mean heterozygosity rate are removed. |
| --pca | | Extracts the top 20 principal components of the variance-standardized relationship matrix. |
| **Default parameter used in PRScs** | | |
| **Parameter** | **Threshold** | **Description** |
| --a | 1 | PARAM_A |
| --b | 0.5 | PARAM_B |
| --phi | 0.01 | PARAM_PHI |
| --n_iter | 1000 | PARAM_ITERATIONS |
| --n_burnin | 500 | PARAM_BURNIN |
| --thin | 5 | PARAM_THINNING_FACTOR |

***Table S3.*** *Features used for the machine learning models, both non-polygenic scores (PGS) and PGS variables.*

| **Non-genetic variables (non-PGS)** | |
|---|---|
| **Variable** | **Description** |
| Sex_Code | Sex |
| Birthmonth | Birth month |
| Somatiska_faktor_Ja_Nej_0 | Presence of somatic disorder(s) (Yes/No) |
| Somatiska_Grav_0 | Severe somatic disorders |
| Somatiska_Hjart_0 | Cardiovascular disorders |
| Somatiska_Endokrina_0 | Endocrine disorders |
| Somatiska_AnnanNeur_0 | Other neurological disorders |
| Somatiska_Annan_0 | Other somatic disorders |
| NeuropsykiatriskHinder_JaNej0 | Autism, learning, coordination or other neurodevelopmental condition |
| NeuropsykiatriskAutism_0 | Autism |
| IQ_group_0 | IQ by group (108-131,93-107,69-92, difficult to assess) |
| Psykos | Hallucination symptoms |
| Tertile_birthmonth | Tertile of birth: January-April, May-August, September-December. |
| Mott_Code | Child & Adolescent Psychiatry unit code |
| Medicationmonth | Month of ADHD medication prescription |
| puls_0 | Heart rate, bpm |
| Blodtryck_Systoliskt_Övre_0 | Systolic blood pressure (upper) |
| Blodtryck_Diastolisk_Undre_0 | Systolic blood pressure (lower) |
| Vikt_0 | Weight |
| Längd_0 | Length |
| CGAS_0 | CGAS total score (baseline) |
| Age_Start | Age at study inclusion |
| Total_SNAP_0 | Total SNAP scores, whole scale (baseline) |
| SNAP_inatt_0 | SNAP inattention subscale scores (baseline) |
| SNAP_hyp_0 | SNAP hyperactivity/impulsivity subscale scores (baseline) |
| SNAP_ODD_0 | SNAP oppositional defiant disorder subscale scores (baseline) |
| SNAP_comb_0 | SNAP subscale combination, inattention and hyperactivity/impulsivity subscale scores (baseline) |
| ASSQ_Tot_0 | ASSQ total scores (baseline) |
| Scas_Tot_0 | SCAS total scores (baseline) |
| MOB_Tot_0 | P-SEC scale total scores (baseline) |

| | |
|---|---|
| interaction | Interaction term SNAP, ASSQ, P-SEC, SCAS and age (baseline) |
| **Genetic variables (PGS)** | |
| **Variable** | **Description** |
| ADHD | ADHD PGS |
| Anxiety | Anxiety PGS |
| ASD | Autism PGS |
| Depression | Depression PGS |
| EA | Educational attainment PGS |
| Intelligence | Intelligence PGS |
| Agreeableness | Agreeableness PGS |
| Conscientiousness | Conscientiousness PGS |
| Extraversion | Extraversion PGS |
| Neuroticism | Neuroticism PGS |
| Openness | Openness PGS |
| PTSD | PTSD PGS |
| Insomnia | Insomnia PGS |
| SleepShort | Short sleep PGS |
| SleepLong | Long sleep PGS |
| Circadian | Circadian rhythm PGS |

***Table S4.*** *Group differences in the secondary clinical variables between individuals with genotyped samples that passed the QC and the remaining participants in the cohort.*

| **Variable** | **Genotyped** | **Not genotyped** | ***p*-value** |
|---|---|---|---|
| SNAP-IV ratio month 1 / baseline | 0.84 ± 0.03 | 0.78 ± 0.02 | 0.161 |
| SNAP-IV ratio month 3 / baseline | 0.79 ± 0.04 | 0.73 ± 0.02 | 0.175 |
| SCAS total (baseline) | 22.95 ± 1.51 | 21.15 ± 0.72 | 0.284 |
| SCAS total (month 3) | 18.94 ± 1.25 | 17.20 ± 0.70 | 0.228 |
| CGAS (baseline) | 50.63 ± 0.69 | 51.16 ± 0.43 | 0.517 |
| CGAS (month 12) | 43.80 ± 12.41 | 52.06 ± 1.55 | 0.496 |
| P-SEC total (baseline) | 19.45 ± 1.13 | 18.93 ± 0.68 | 0.690 |
| Weight (baseline) | 48.74 ± 2.83 | 49.22 ± 2.07 | 0.892 |
| Height (baseline) | 151.94 ± 1.92 | 151.66 ± 1.07 | 0.900 |
| Heart rate (baseline) | 75.56 ± 1.03 | 75.48 ± 0.64 | 0.948 |
| Systolic blood pressure (baseline) | 109.72 ± 1.03 | 111.02 ± 0.71 | 0.302 |
| Diastolic blood pressure (baseline) | 66.14 ± 0.79 | 65.58 ± 0.45 | 0.540 |
| Month of birth | | | 0.195 |
| • January | 37 (12%) | 6 (5.5%) | |
| • February | 27 (8.7%) | 5 (4.5%) | |
| • March | 21 (6.8%) | 11 (10%) | |
| • April | 24 (7.8%) | 10 (9.1%) | |
| • May | 22 (7.1%) | 7 (6.4%) | |
| • June | 19 (6.1%) | 7 (6.4%) | |
| • July | 24 (7.8%) | 10 (9.1%) | |
| • August | 31 (10%) | 5 (4.5%) | |
| • September | 20 (6.5%) | 9 (8.2%) | |
| • October | 32 (10.4%) | 11 (10%) | |
| • November | 27 (8) | 12 (10.9%) | |
| • December | 25 (8.1%) | 17 (15.5%) | |
| Tertile of birth | | | 0.125 |
| • January – April | 109 (35.3%) | 32 (29.1%) | |
| • May – August | 96 (31.1%) | 29 (26.4%) | |
| • September – December | 104 (33.7%) | 49 (44.5%) | |
| **Child- and Adolescent Units (CAPs)** | | | **5.00E-04** |

| | | | |
|---|---|---|---|
| • Brommaplan | 5 (1.6%) | 6 (5.5%) | |
| • Danderyd | 36 (11.7%) | 38 (34.5%) | |
| • Ektorp | 51 (16.5%) | 10 (9.1%) | |
| • Farsta | 9 (2.9%) | 11 (10%) | |
| • Globen | 28 (9.1%) | 14 (12.7%) | |
| • Gotland | 16 (5.2%) | 2 (1.8%) | |
| • Huddinge | 5 (1.6%) | 2 (1.8%) | |
| • Kungsholmen | 7 (2.3%) | 0 (0%) | |
| • OCD | 1 (0.3%) | 0 (0%) | |
| • Skärholmen | 39 (12.6%) | 13 (11.8%) | |
| • Sodertälje | 11 (3.6%) | 1 (0.9%) | |
| • Sollentuna | 4 (1.3%) | 4 (3.6%) | |
| • Solna | 11 (3.6%) | 5 (4.5%) | |
| • Umeå | 86 (27.8%) | 4 (3.6%) | |
| **CGAS groups (baseline)** | | | **0.043** |
| • Severe problems (21-30) | 1 (0.4%) | 0 (0%) | |
| • Serious problems (31-40) | 10 (4.1%) | 6 (7.6%) | |
| • Obvious problems (41-50) | 100 (40.7%) | 30 (38%) | |
| • Some noticeable problems (51-60) | 118 (48%) | 43 (54.4%) | |
| • Some problems (61-70) | 17 (6.9%) | 0 (0%) | |
| Medication ADHD (No) | 33 (13%) | 4 (6%) | 0.166 |
| Medication month | | | 0.287 |
| • January | 27 (8.7%) | 7 (6.4%) | |
| • February | 20 (6.5%) | 11 (10%) | |
| • March | 22 (7.1%) | 9 (8.2%) | |
| • April | 20 (6.5%) | 10 (9.1%) | |
| • May | 27 (8.7%) | 10 (9.1%) | |
| • June | 26 (8.4%) | 2 (1.8%) | |
| • July | 5 (1.6%) | 5 (4.5%) | |
| • August | 37 (12%) | 11 (10%) | |
| • September | 33 (10.7%) | 15 (13.6%) | |
| • October | 40 (12.9%) | 12 (10.9%) | |
| • November | 33 (10.7%) | 11 (10%) | |

| | | | |
|---|---|---|---|
| • December | 19 (6.1%) | 7 (6.4%) | |
| Medication tertile | | | 0.495 |
| • January – April | 89 (28.8%) | 37 (33.6%) | |
| • May – August | 95 (30.7%) | 28 (25.5%) | |
| • September – December | 125 (40.5%) | 45 (40.9%) | |
| Responder inattention (month 3) | | | 0.389 |
| • Non-responder | 120 (39.1%) | 50 (45.5%) | |
| • Intermediate responder | 77 (25.1%) | 28 (25.5%) | |
| • Responder | 110 (35.8%) | 32 (29.1%) | |
| Responder hyperactivity (month 3) | | | 0.677 |
| • Non-responder | 107 (35.1%) | 43 (39.8%) | |
| • Intermediate responder | 62 (20.3%) | 20 (18.5%) | |
| • Responder | 136 (44.6%) | 45 (41.7%) | |
| Responder SNAP-IV odd (month 3) | | | 0.732 |
| • Non-responder | 147 (47.6%) | 51 (46.4%) | |
| • Intermediate responder | 49 (15.9%) | 21 (19.1%) | |
| • Responder | 113 (36.6%) | 38 (34.5%) | |

*For continuous variables, values are presented as mean ± standard error of the mean (SEM). Normality was evaluated separately within each group using the Shapiro-Wilk test. Group comparisons were performed using Student's t-test for normally distributed variables or the Wilcoxon rank-sum test otherwise. For categorical variables, data are presented as counts and percentages. Group differences were evaluated using the chi-square test of independence. When expected cell counts were below five, Fisher's exact test was applied. All statistical tests were two-sided, and p-values < 0.05 were considered statistically significant.*

***Table S5.*** *Associations between polygenic scores (PGS) and Swanson, Nolan, and Pelham ADHD Rating Scale-version IV (SNAP-IV) scores.*

| | Entire cohort | | | | Age ≥ 13 | | | | Age < 13 | | | |
|---|---|---|---|---|---|---|---|---|---|---|---|---|
| | ***β*** | **SE** | **p-value** | **p FDR** | ***β*** | **SE** | **p-value** | **p FDR** | ***β*** | **SE** | **p-value** | **p FDR** |
| **ADHD** | | | | | | | | | | | | |
| ADHD PGS | 0.1419 | 0.0531 | 7.82E-03 | 2.93E-02 | 0.2319 | 0.0876 | 8.97E-03 | 3.59E-02 | 0.0857 | 0.0685 | 2.11E-01 | 4.61E-01 |
| Time*ADHD PGS | -0.0146 | 0.0474 | 7.58E-01 | 7.93E-01 | -0.0467 | 0.071 | 5.13E-01 | 5.13E-01 | 0.0138 | 0.0631 | 8.27E-01 | 8.94E-01 |
| Time | -0.7542 | 0.0473 | 3.78E-42 | 3.47E-41 | -0.6731 | 0.0706 | 2.45E-16 | 1.96E-15 | -0.81 | 0.0632 | 2.26E-27 | 1.81E-26 |
| Age | -0.2237 | 0.0471 | 3.13E-06 | 1.56E-05 | | | | | | | | |
| Sex | -0.0427 | 0.0982 | 6.64E-01 | 7.12E-01 | -0.2152 | 0.1461 | 1.43E-01 | 3.04E-01 | 0.1435 | 0.1347 | 2.88E-01 | 4.61E-01 |
| PC1 | 0.0801 | 0.0696 | 2.51E-01 | 4.18E-01 | 0.1741 | 0.1207 | 1.52E-01 | 3.04E-01 | 0.0119 | 0.0894 | 8.94E-01 | 8.94E-01 |
| PC2 | 0.1313 | 0.0814 | 1.08E-01 | 2.11E-01 | 0.1146 | 0.1084 | 2.93E-01 | 3.90E-01 | 0.1452 | 0.1214 | 2.33E-01 | 4.61E-01 |
| PC3 | -0.0601 | 0.0831 | 4.71E-01 | 5.46E-01 | -0.1523 | 0.1178 | 1.99E-01 | 3.18E-01 | 0.0325 | 0.118 | 7.83E-01 | 8.94E-01 |
| PC4 | 0.1642 | 0.0666 | 1.42E-02 | 3.99E-02 | 0.0783 | 0.0948 | 4.11E-01 | 4.69E-01 | 0.1986 | 0.0966 | 4.12E-02 | 1.65E-01 |
| **Autism** | | | | | | | | | | | | |
| Autism PGS | 0.0415 | 0.0517 | 4.23E-01 | 5.28E-01 | 0.0864 | 0.0791 | 2.77E-01 | 3.66E-01 | -0.0251 | 0.0693 | 7.17E-01 | 9.21E-01 |
| Time*Autism PGS | 0.0058 | 0.0474 | 9.02E-01 | 9.02E-01 | 0.0607 | 0.0669 | 3.66E-01 | 3.66E-01 | -0.0311 | 0.0655 | 6.36E-01 | 9.21E-01 |
| Time | -0.7542 | 0.0473 | 3.85E-42 | 3.47E-41 | -0.6656 | 0.0703 | 3.41E-16 | 2.73E-15 | -0.8082 | 0.0631 | 2.40E-27 | 1.92E-26 |
| Age | -0.2328 | 0.0475 | 1.57E-06 | 1.18E-05 | | | | | | | | |
| Sex | -0.0655 | 0.0992 | 5.10E-01 | 5.74E-01 | -0.1904 | 0.1519 | 2.12E-01 | 3.66E-01 | 0.1078 | 0.1339 | 4.22E-01 | 8.44E-01 |
| PC1 | 0.065 | 0.0702 | 3.55E-01 | 5.06E-01 | 0.1383 | 0.1215 | 2.57E-01 | 3.66E-01 | -0.0023 | 0.0898 | 9.79E-01 | 9.79E-01 |
| PC2 | 0.1449 | 0.0822 | 7.93E-02 | 1.70E-01 | 0.1266 | 0.1102 | 2.53E-01 | 3.66E-01 | 0.1603 | 0.122 | 1.91E-01 | 5.08E-01 |
| PC3 | -0.0773 | 0.0838 | 3.58E-01 | 5.06E-01 | -0.1941 | 0.12 | 1.09E-01 | 3.66E-01 | 0.0292 | 0.1187 | 8.06E-01 | 9.21E-01 |
| PC4 | 0.1676 | 0.0674 | 1.34E-02 | 3.99E-02 | 0.0907 | 0.0963 | 3.48E-01 | 3.66E-01 | 0.2051 | 0.0973 | 3.63E-02 | 1.45E-01 |
| **EA** | | | | | | | | | | | | |
| EA PGS | -0.1161 | 0.0519 | 2.56E-02 | 6.79E-02 | -0.1488 | 0.0896 | 9.85E-02 | 2.01E-01 | -0.0958 | 0.0649 | 1.41E-01 | 3.77E-01 |
| Time*EA PGS | 0.0946 | 0.0471 | 4.53E-02 | 1.13E-01 | 0.163 | 0.075 | 3.18E-02 | 1.27E-01 | 0.0528 | 0.0605 | 3.84E-01 | 5.50E-01 |
| Time | -0.7542 | 0.047 | 1.55E-42 | 3.47E-41 | -0.6846 | 0.0695 | 4.16E-17 | 3.33E-16 | -0.8059 | 0.0631 | 3.00E-27 | 2.40E-26 |
| Age | -0.2297 | 0.0475 | 2.11E-06 | 1.18E-05 | | | | | | | | |
| Sex | -0.0708 | 0.0985 | 4.73E-01 | 5.46E-01 | -0.2316 | 0.1502 | 1.26E-01 | 2.01E-01 | 0.1095 | 0.1334 | 4.13E-01 | 5.50E-01 |
| PC1 | 0.0627 | 0.0698 | 3.70E-01 | 5.06E-01 | 0.1239 | 0.1221 | 3.12E-01 | 3.12E-01 | 0.0082 | 0.0895 | 9.27E-01 | 9.33E-01 |
| PC2 | 0.1496 | 0.0816 | 6.84E-02 | 1.59E-01 | 0.1437 | 0.1107 | 1.97E-01 | 2.62E-01 | 0.1439 | 0.1219 | 2.39E-01 | 4.79E-01 |
| PC3 | -0.0786 | 0.0833 | 3.48E-01 | 5.06E-01 | -0.1885 | 0.1215 | 1.23E-01 | 2.01E-01 | 0.0101 | 0.1188 | 9.33E-01 | 9.33E-01 |
| PC4 | 0.185 | 0.0676 | 6.58E-03 | 2.69E-02 | 0.1084 | 0.0978 | 2.70E-01 | 3.09E-01 | 0.2131 | 0.0973 | 2.98E-02 | 1.19E-01 |

| | | | | | | | | | | | | |
|---|---|---|---|---|---|---|---|---|---|---|---|---|
| **Intelligence** | | | | | | | | | | | | |
| Intelligence PGS | -0.0518 | 0.0528 | 3.28E-01 | 5.06E-01 | -0.0638 | 0.0905 | 4.82E-01 | 4.82E-01 | -0.0399 | 0.0662 | 5.47E-01 | 8.76E-01 |
| Time*Intelligence PGS | 0.06 | 0.0472 | 2.05E-01 | 3.55E-01 | 0.1617 | 0.0726 | 2.78E-02 | 1.11E-01 | -0.004 | 0.0616 | 9.48E-01 | 9.89E-01 |
| Time | -0.7542 | 0.0472 | 2.68E-42 | 3.47E-41 | -0.6838 | 0.0694 | 4.04E-17 | 3.23E-16 | -0.8094 | 0.0632 | 2.42E-27 | 1.94E-26 |
| Age | -0.232 | 0.0477 | 1.83E-06 | 1.18E-05 | | | | | | | | |
| Sex | -0.0767 | 0.0988 | 4.38E-01 | 5.33E-01 | -0.2464 | 0.1503 | 1.04E-01 | 2.77E-01 | 0.1036 | 0.1342 | 4.41E-01 | 8.76E-01 |
| PC1 | 0.0572 | 0.0701 | 4.16E-01 | 5.28E-01 | 0.1181 | 0.1224 | 3.37E-01 | 3.97E-01 | -0.0012 | 0.0897 | 9.89E-01 | 9.89E-01 |
| PC2 | 0.1491 | 0.0821 | 7.08E-02 | 1.59E-01 | 0.1429 | 0.111 | 2.01E-01 | 3.21E-01 | 0.15 | 0.122 | 2.21E-01 | 5.88E-01 |
| PC3 | -0.0756 | 0.0842 | 3.71E-01 | 5.06E-01 | -0.1722 | 0.1224 | 1.62E-01 | 3.21E-01 | 0.0179 | 0.1188 | 8.81E-01 | 9.89E-01 |
| PC4 | 0.1753 | 0.0679 | 1.03E-02 | 3.30E-02 | 0.0937 | 0.0993 | 3.47E-01 | 3.97E-01 | 0.2065 | 0.0973 | 3.52E-02 | 1.41E-01 |
| **PTSD** | | | | | | | | | | | | |
| PTSD PGS | 0.0872 | 0.0649 | 1.80E-01 | 3.24E-01 | 0.252 | 0.1146 | 2.94E-02 | 1.18E-01 | 0.0223 | 0.0794 | 7.79E-01 | 8.57E-01 |
| Time*PTSD PGS | 0.0079 | 0.0474 | 8.68E-01 | 8.88E-01 | 5.00E-04 | 0.0762 | 9.95E-01 | 9.95E-01 | 0.0192 | 0.0607 | 7.52E-01 | 8.57E-01 |
| Time | -0.7542 | 0.0473 | 3.84E-42 | 3.47E-41 | -0.6687 | 0.0708 | 3.83E-16 | 3.06E-15 | -0.8103 | 0.0632 | 2.13E-27 | 1.70E-26 |
| Age | -0.2229 | 0.0479 | 4.92E-06 | 2.21E-05 | | | | | | | | |
| Sex | -0.0494 | 0.1 | 6.21E-01 | 6.82E-01 | -0.1581 | 0.151 | 2.97E-01 | 3.43E-01 | 0.1187 | 0.1348 | 3.80E-01 | 7.59E-01 |
| PC1 | 0.1045 | 0.0758 | 1.69E-01 | 3.17E-01 | 0.2429 | 0.131 | 6.64E-02 | 1.77E-01 | 0.0174 | 0.0965 | 8.57E-01 | 8.57E-01 |
| PC2 | 0.1368 | 0.0823 | 9.78E-02 | 2.00E-01 | 0.1137 | 0.1091 | 3.00E-01 | 3.43E-01 | 0.1516 | 0.1222 | 2.16E-01 | 5.77E-01 |
| PC3 | -0.0676 | 0.0838 | 4.22E-01 | 5.28E-01 | -0.1613 | 0.1183 | 1.75E-01 | 3.43E-01 | 0.0278 | 0.1188 | 8.15E-01 | 8.57E-01 |
| PC4 | 0.176 | 0.0673 | 9.34E-03 | 3.23E-02 | 0.1145 | 0.0953 | 2.32E-01 | 3.43E-01 | 0.2032 | 0.0973 | 3.80E-02 | 1.52E-01 |

*Linear mixed-effects models tested associations between standardized polygenic scores (PGS) and Swanson, Nolan, and Pelham ADHD Rating Scale-version IV (SNAP-IV) scores at baseline and 3 months, including PGS main effects and PGS x Time interactions, adjusted for age (not in age-stratified analyses), sex, and the first four principal components (PC), with random intercepts for participant and region. Estimates are shown as β, standard error (SE), nominal p-values and FDR adjusted p-values. Models are reported for each trait-specific PGS, including fixing effects and interaction terms. Columns are presented for the entire cohort and stratified by age group (≥13 and <13 years).*

***Table S6.*** *Associations between polygenic scores (PGS) and Swanson, Nolan, and Pelham ADHD Rating Scale-version IV (SNAP-IV) scores.*

| | Responder | | | | Intermediate responder | | | | Non-responder | | | |
|---|---|---|---|---|---|---|---|---|---|---|---|---|
| | ***β*** | **SE** | **p-value** | **p FDR** | ***β*** | **SE** | **p-value** | **p FDR** | ***β*** | **SE** | **p-value** | **p FDR** |
| **ADHD** | | | | | | | | | | | | |
| ADHD PGS | 0.14 | 0.0736 | 5.93E-02 | 1.48E-01 | 0.1472 | 0.0901 | 1.06E-01 | 3.60E-01 | 0.1604 | 0.0873 | 6.86E-02 | 5.15E-01 |
| Time*ADHD PGS | -0.1084 | 0.0677 | 1.12E-01 | 2.11E-01 | -0.0214 | 0.0331 | 5.19E-01 | 8.22E-01 | 0.0137 | 0.0394 | 7.28E-01 | 9.36E-01 |
| Time | -1.5764 | 0.0684 | 8.51E-42 | 1.91E-40 | -0.8047 | 0.0319 | 8.12E-42 | 1.22E-40 | -0.0105 | 0.04 | 7.93E-01 | 9.36E-01 |
| Age | -0.2672 | 0.0717 | 3.31E-04 | 1.66E-03 | -0.2268 | 0.0864 | 1.03E-02 | 4.65E-02 | -0.2138 | 0.0799 | 8.60E-03 | 1.15E-01 |
| Sex | -0.0393 | 0.1357 | 7.73E-01 | 7.90E-01 | -0.068 | 0.1798 | 7.06E-01 | 8.22E-01 | -0.1199 | 0.1813 | 5.10E-01 | 8.82E-01 |
| PC1 | 0.1995 | 0.1389 | 1.54E-01 | 2.33E-01 | 0.0651 | 0.1389 | 6.41E-01 | 8.22E-01 | 0.0834 | 0.108 | 4.41E-01 | 8.28E-01 |
| PC2 | 0.3932 | 0.1684 | 2.17E-02 | 6.65E-02 | 0.0322 | 0.1225 | 7.93E-01 | 8.30E-01 | 0.1409 | 0.1427 | 3.30E-01 | 7.20E-01 |
| PC3 | 0.1125 | 0.1444 | 4.38E-01 | 5.26E-01 | -0.2706 | 0.1656 | 1.06E-01 | 3.60E-01 | 0.0207 | 0.1541 | 8.94E-01 | 9.58E-01 |
| PC4 | 0.2517 | 0.1458 | 8.76E-02 | 2.08E-01 | -0.2486 | 0.248 | 3.19E-01 | 6.66E-01 | 0.111 | 0.105 | 2.93E-01 | 7.20E-01 |
| **Autism** | | | | | | | | | | | | |
| Autism PGS | 0.1032 | 0.0664 | 1.23E-01 | 2.21E-01 | 0.0909 | 0.0919 | 3.26E-01 | 6.66E-01 | -0.063 | 0.0902 | 4.86E-01 | 8.76E-01 |
| Time*Autism PGS | -0.1325 | 0.0621 | 3.52E-02 | 9.91E-02 | -0.0281 | 0.0345 | 4.18E-01 | 7.83E-01 | 0.0235 | 0.042 | 5.77E-01 | 9.36E-01 |
| Time | -1.5925 | 0.0684 | 3.75E-42 | 1.69E-40 | -0.8017 | 0.0321 | 1.94E-41 | 1.75E-40 | -0.0113 | 0.04 | 7.79E-01 | 9.36E-01 |
| Age | -0.2695 | 0.0722 | 3.22E-04 | 1.66E-03 | -0.2409 | 0.0867 | 6.75E-03 | 3.37E-02 | -0.223 | 0.081 | 6.88E-03 | 1.15E-01 |
| Sex | -0.0398 | 0.1367 | 7.72E-01 | 7.90E-01 | -0.0779 | 0.184 | 6.73E-01 | 8.22E-01 | -0.2025 | 0.182 | 2.68E-01 | 7.20E-01 |
| PC1 | 0.2025 | 0.1398 | 1.51E-01 | 2.33E-01 | 0.0248 | 0.1388 | 8.58E-01 | 8.78E-01 | 0.0308 | 0.1125 | 7.85E-01 | 9.36E-01 |
| PC2 | 0.4066 | 0.1692 | 1.82E-02 | 6.65E-02 | 0.0382 | 0.1239 | 7.58E-01 | 8.22E-01 | 0.1674 | 0.1447 | 2.56E-01 | 7.20E-01 |
| PC3 | 0.1152 | 0.1454 | 4.30E-01 | 5.26E-01 | -0.2545 | 0.1672 | 1.32E-01 | 3.71E-01 | 0.0322 | 0.1598 | 8.42E-01 | 9.47E-01 |
| PC4 | 0.2426 | 0.1467 | 1.02E-01 | 2.08E-01 | -0.2031 | 0.2479 | 4.15E-01 | 7.83E-01 | 0.1146 | 0.1064 | 2.85E-01 | 7.20E-01 |
| **EA** | | | | | | | | | | | | |
| EA PGS | -0.0907 | 0.0753 | 2.31E-01 | 3.24E-01 | -0.114 | 0.0776 | 1.45E-01 | 3.85E-01 | -0.1159 | 0.0908 | 2.04E-01 | 7.20E-01 |
| Time*EA PGS | 0.1391 | 0.069 | 4.66E-02 | 1.23E-01 | 0.0413 | 0.0296 | 1.67E-01 | 4.11E-01 | -0.017 | 0.0432 | 6.94E-01 | 9.36E-01 |
| Time | -1.5454 | 0.0689 | 8.67E-41 | 8.16E-40 | -0.8144 | 0.0322 | 8.11E-42 | 1.22E-40 | -0.0103 | 0.04 | 7.97E-01 | 9.36E-01 |
| Age | -0.2694 | 0.0723 | 3.30E-04 | 1.66E-03 | -0.2555 | 0.085 | 3.54E-03 | 2.60E-02 | -0.2057 | 0.0813 | 1.28E-02 | 1.15E-01 |
| Sex | -0.0426 | 0.1369 | 7.56E-01 | 7.90E-01 | -0.0917 | 0.1792 | 6.10E-01 | 8.22E-01 | -0.1922 | 0.1794 | 2.86E-01 | 7.20E-01 |
| PC1 | 0.2009 | 0.14 | 1.55E-01 | 2.33E-01 | 0.0411 | 0.1381 | 7.67E-01 | 8.22E-01 | 0.0495 | 0.1072 | 6.45E-01 | 9.36E-01 |
| PC2 | 0.4008 | 0.1723 | 2.22E-02 | 6.65E-02 | 0.0507 | 0.123 | 6.81E-01 | 8.22E-01 | 0.161 | 0.141 | 2.56E-01 | 7.20E-01 |
| PC3 | 0.1124 | 0.1464 | 4.45E-01 | 5.26E-01 | -0.2749 | 0.1666 | 1.03E-01 | 3.60E-01 | 0.0079 | 0.152 | 9.58E-01 | 9.58E-01 |
| PC4 | 0.2402 | 0.1472 | 1.06E-01 | 2.08E-01 | -0.1348 | 0.2471 | 5.87E-01 | 8.22E-01 | 0.1362 | 0.1059 | 2.01E-01 | 7.20E-01 |

| | | | | | | | | | | | | |
|---|---|---|---|---|---|---|---|---|---|---|---|---|
| **Intelligence** | | | | | | | | | | | | |
| Intelligence PGS | -0.0451 | 0.0807 | 5.77E-01 | 6.49E-01 | -0.0626 | 0.0815 | 4.44E-01 | 8.00E-01 | -0.0334 | 0.086 | 6.99E-01 | 9.36E-01 |
| Time*Intelligence PGS | 0.0942 | 0.0748 | 2.11E-01 | 3.06E-01 | -0.0101 | 0.0305 | 7.41E-01 | 8.22E-01 | -0.0208 | 0.0394 | 5.99E-01 | 9.36E-01 |
| Time | -1.5568 | 0.0695 | 9.06E-41 | 8.16E-40 | -0.804 | 0.0322 | 1.73E-41 | 1.75E-40 | -0.0096 | 0.0399 | 8.11E-01 | 9.36E-01 |
| Age | -0.2695 | 0.0723 | 3.33E-04 | 1.66E-03 | -0.251 | 0.0855 | 4.34E-03 | 2.60E-02 | -0.2179 | 0.0821 | 9.12E-03 | 1.15E-01 |
| Sex | -0.0435 | 0.137 | 7.52E-01 | 7.90E-01 | -0.1128 | 0.1792 | 5.31E-01 | 8.22E-01 | -0.1926 | 0.1807 | 2.89E-01 | 7.20E-01 |
| PC1 | 0.2017 | 0.1408 | 1.55E-01 | 2.33E-01 | 0.0105 | 0.1408 | 9.41E-01 | 9.41E-01 | 0.0543 | 0.1087 | 6.19E-01 | 9.36E-01 |
| PC2 | 0.4115 | 0.1705 | 1.77E-02 | 6.65E-02 | 0.0417 | 0.1234 | 7.36E-01 | 8.22E-01 | 0.1495 | 0.1448 | 3.10E-01 | 7.20E-01 |
| PC3 | 0.1174 | 0.1458 | 4.22E-01 | 5.26E-01 | -0.2689 | 0.1674 | 1.12E-01 | 3.60E-01 | -0.012 | 0.1594 | 9.41E-01 | 9.58E-01 |
| PC4 | 0.2432 | 0.147 | 1.01E-01 | 2.08E-01 | -0.1614 | 0.2466 | 5.15E-01 | 8.22E-01 | 0.1205 | 0.1075 | 2.66E-01 | 7.20E-01 |
| **PTSD** | | | | | | | | | | | | |
| PTSD PGS | 0.0585 | 0.0929 | 5.31E-01 | 6.12E-01 | 0.112 | 0.1058 | 2.93E-01 | 6.59E-01 | 0.1131 | 0.1138 | 3.22E-01 | 7.20E-01 |
| Time*PTSD PGS | -0.0767 | 0.0694 | 2.72E-01 | 3.70E-01 | -0.0507 | 0.0328 | 1.26E-01 | 3.71E-01 | 0.0022 | 0.0389 | 9.55E-01 | 9.58E-01 |
| Time | -1.577 | 0.069 | 1.73E-41 | 2.60E-40 | -0.8056 | 0.0315 | 3.08E-42 | 1.22E-40 | -0.0103 | 0.0401 | 7.98E-01 | 9.36E-01 |
| Age | -0.2664 | 0.0735 | 4.71E-04 | 2.12E-03 | -0.2493 | 0.0856 | 4.63E-03 | 2.60E-02 | -0.2096 | 0.0821 | 1.21E-02 | 1.15E-01 |
| Sex | -0.0361 | 0.1406 | 7.98E-01 | 7.98E-01 | -0.0873 | 0.182 | 6.33E-01 | 8.22E-01 | -0.1707 | 0.181 | 3.48E-01 | 7.20E-01 |
| PC1 | 0.2101 | 0.145 | 1.51E-01 | 2.33E-01 | 0.0607 | 0.143 | 6.72E-01 | 8.22E-01 | 0.1189 | 0.127 | 3.52E-01 | 7.20E-01 |
| PC2 | 0.4062 | 0.1706 | 1.92E-02 | 6.65E-02 | 0.0386 | 0.1237 | 7.56E-01 | 8.22E-01 | 0.1275 | 0.1464 | 3.91E-01 | 7.64E-01 |
| PC3 | 0.1193 | 0.1459 | 4.15E-01 | 5.26E-01 | -0.2343 | 0.1707 | 1.74E-01 | 4.11E-01 | -0.0138 | 0.1565 | 9.31E-01 | 9.58E-01 |
| PC4 | 0.2435 | 0.147 | 1.01E-01 | 2.08E-01 | -0.1555 | 0.2475 | 5.31E-01 | 8.22E-01 | 0.1163 | 0.1063 | 2.77E-01 | 7.20E-01 |

*Linear mixed-effects models tested associations between standardized polygenic scores (PGS) and Swanson, Nolan, and Pelham ADHD Rating Scale-version IV (SNAP-IV) scores at baseline and 3 months, including PGS main effects and PGS x Time interactions, adjusted for age (not in age-stratified analyses), sex, and the first four principal components (PC), with random intercepts for participant and region. Estimates are shown as β, standard error (SE), nominal p-values and FDR adjusted p-values. Models are reported for each trait-specific PGS, including fixing effects and interaction terms. Columns are presented for treatment outcome (responders, intermediate responders and non-responders).*

### Supplementary Figures

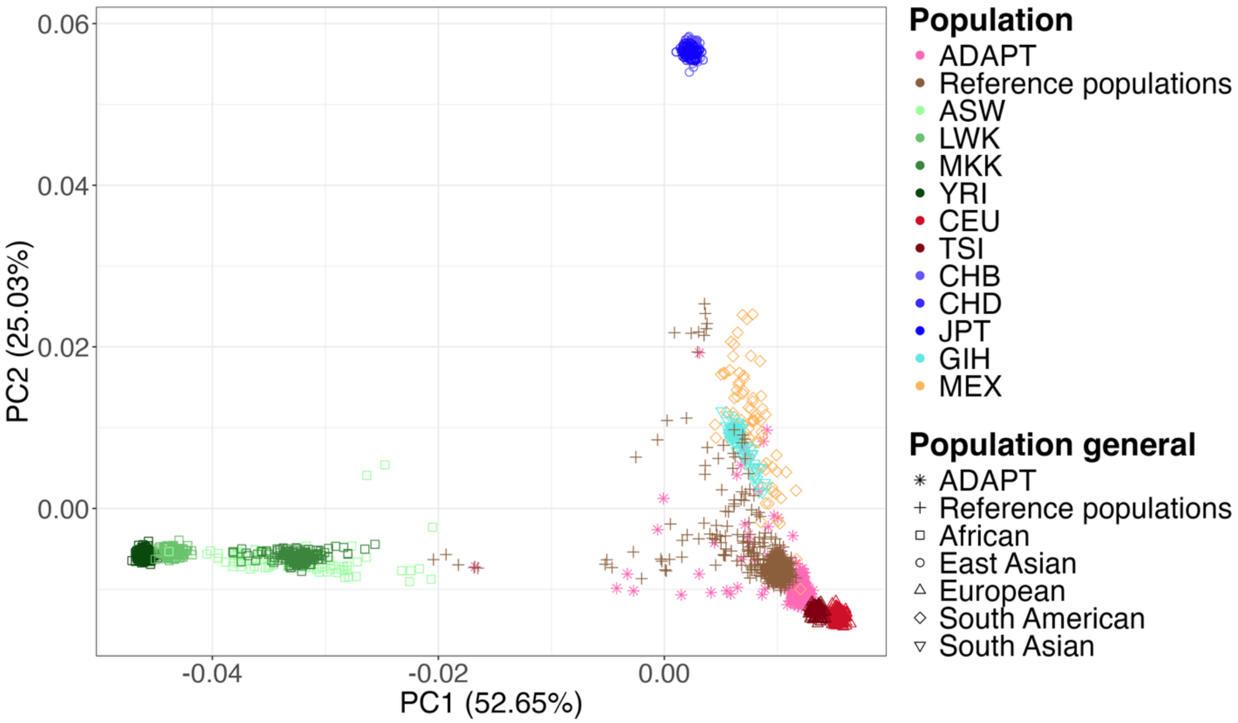

***Figure S1*** *Principal component analysis (PCA) for genetic ancestry estimation, showing Principal Component (PC) 1 and PC2, with the percentage of variance explained by each PC indicated in parenthesis. Shapes denote the general population, and colors indicate HapMap Phase III reference populations. The ADAPT cohort is highlighted in pink with asterisks, and the reference population in brown with crosses*

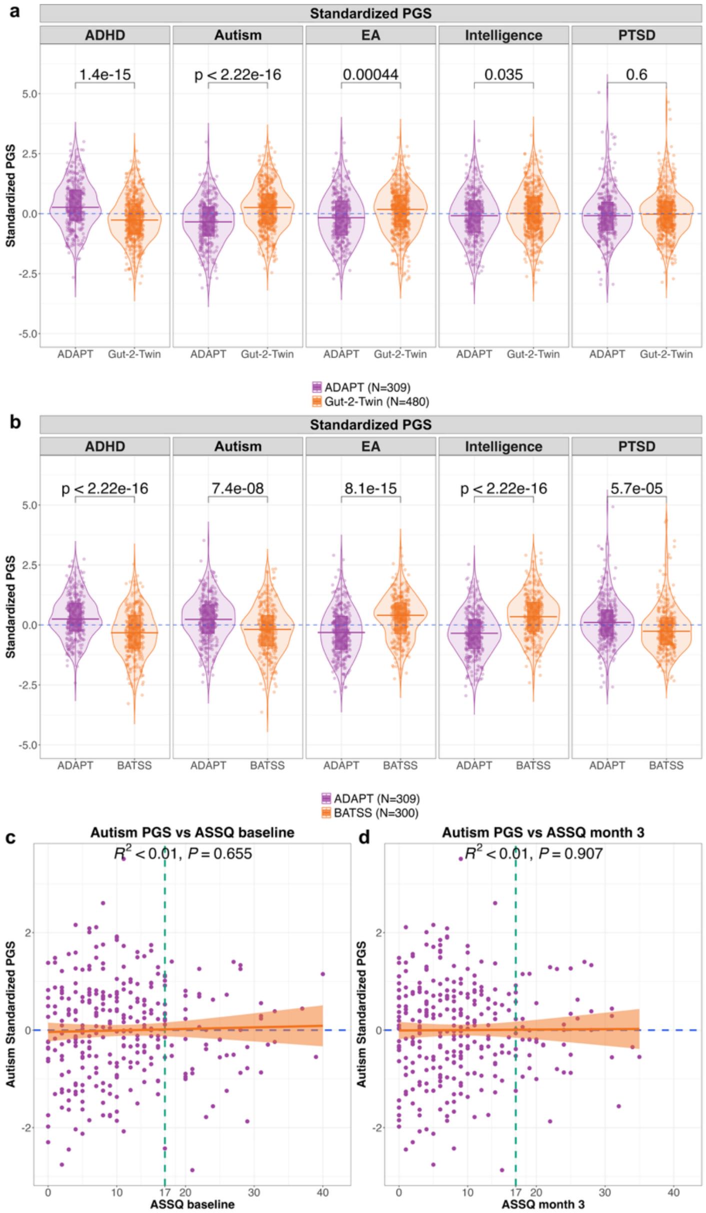

***Figure S2 Polygenic scores (PGS) across cohorts and clinical correlations. A-B)*** *Comparison between ADAPT cohort (N = 309) and the Gut-2-Twin reference cohort (N = 480,* ***A****) and the BATSS reference cohort (N = 300,* ***B****) of the distribution of the different PGS: attention deficit hyperactivity disorder (ADHD), autism, educational attainment (EA), intelligence and posttraumatic stress disorder (PTSD). The p-values from Student's t-test are shown.* ***C-D)*** *Correlation between polygenic scores (PGSs) for autism and Autism Spectrum Screening Questionnaire (ASSQ) at baseline* ***(C)*** *and at 3 months* ***(D)****. Green dashed vertical line indicates the ASSQ cut-off for autism classification, where scores ≥ 17 are considered autistic. Orange lines represent the linear model fitted for each PGS by each clinical variable, and the orange curve shows the 95% confidence interval of the predicted lineal model*

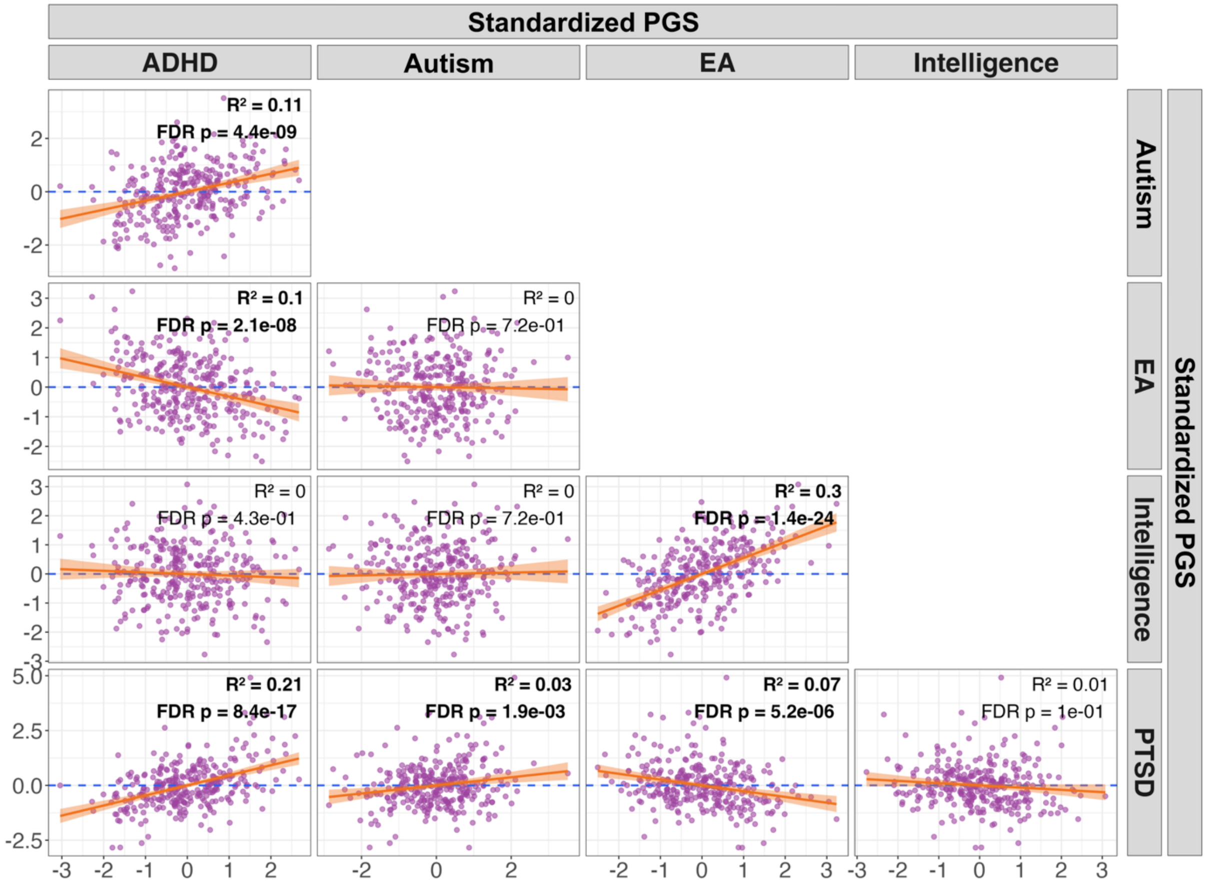

***Figure S3*** *Pairwise correlation plots among standardized polygenic scores (PGSs) for attention deficit hyperactivity disorder (ADHD), autism, educational attainment (EA), intelligence and posttraumatic stress disorder (PTSD). Each panel displays pairwise associations with fitted linear regression lines (orange) and the 95% confidence interval (orange shading), together with the coefficient of determination ($R^2$) and corresponding p-value corrected by False Discovery Rate (FDR). Significant associations are shown in bold*

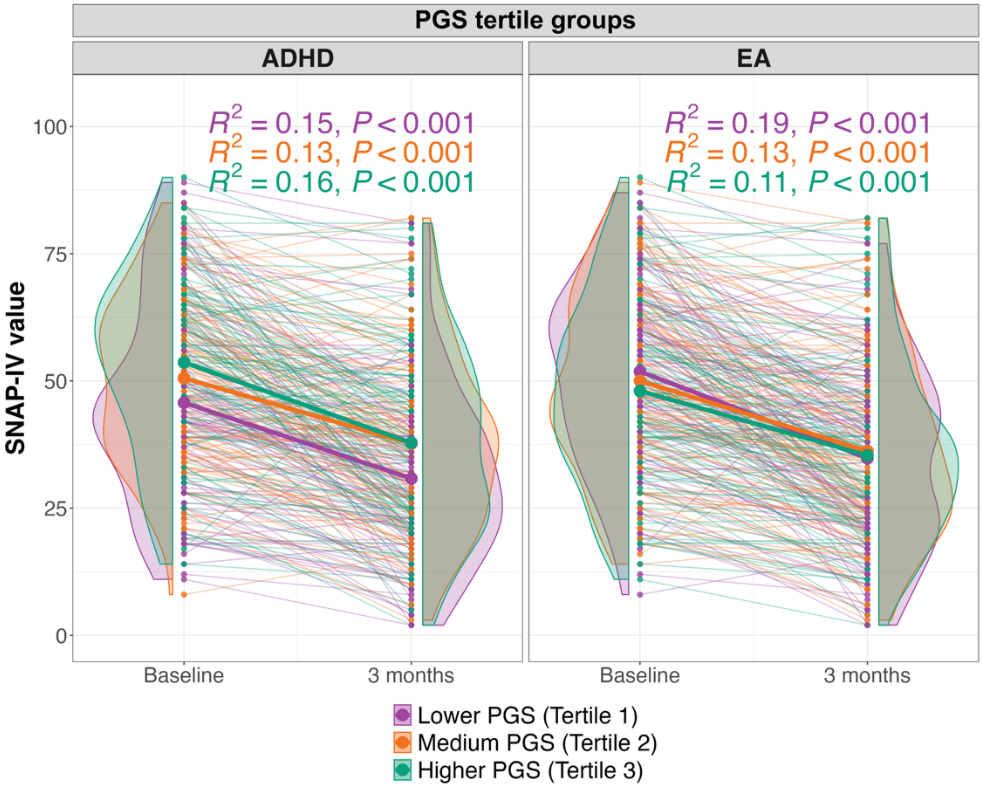

***Figure S4 SNAP-IV trajectories by polygenic scores (PGS) tertiles for ADHD and EA.*** *Longitudinal SNAP-IV scores from baseline to 3-months for individuals stratified by tertiles of PGS for ADHD (left) and EA (right). Individual trajectories, groups means, and score distributions are shown. Colors indicate PGS tertiles*

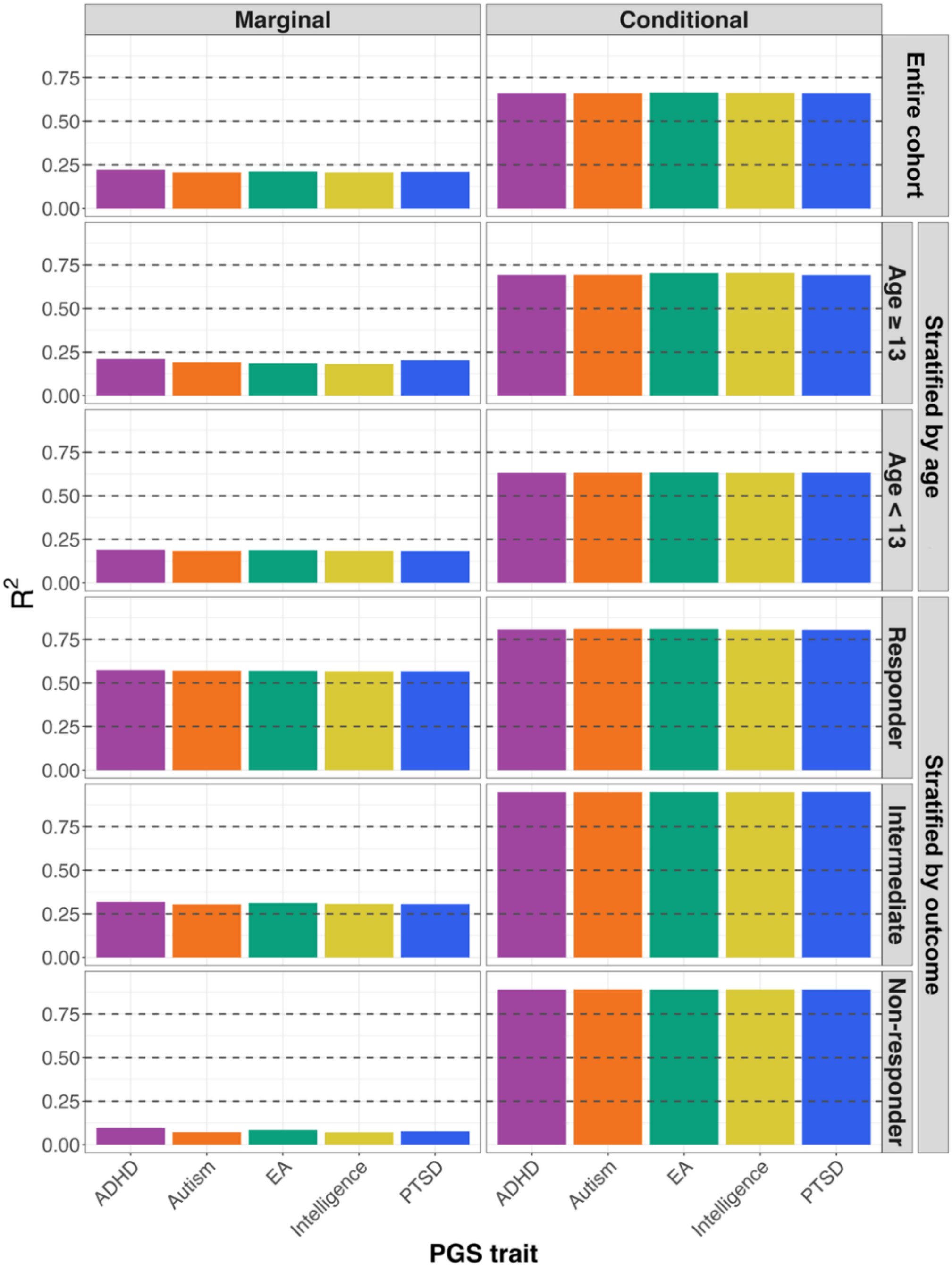

***Figure S5 Variance explained by the associations between polygenic scores (PGS) and Swanson, Nolan, and Pelham ADHD Rating Scale-version IV (SNAP-IV) scores.*** *Linear mixed-effects models tested associations between standardized PGS and SNAP-IV scores over time (baseline vs 3 months). Including PGS main effects and PGS × Time interactions, adjusting for age, sex, and the first four genetic principal components, with random intercepts for participant and region. Marginal and conditional $R^2$ are shown for each trait (by colors) and organized in panels for the entire cohort, by age group (≥13 and <13 years) and by treatment outcome (responders, intermediate responders and non-responders)*
